# Nationwide Organ Volume Reference Standards and Aging-Related Changes in Abdominal CT from Japan

**DOI:** 10.64898/2026.01.30.26345246

**Authors:** Tomohiro Kikuchi, Kohei Yamamoto, Yosuke Yamagishi, Toshiaki Akashi, Shouhei Hanaoka, Takeharu Yoshikawa, Hiroyuki Fujii, Harushi Mori, Hisaki Makimoto, Takahide Kohro

**Affiliations:** Data Science Center, Jichi Medical University, 3311-1 Yakushiji, Shimotsuke, Tochigi, 329-0498, Japan; Department of Radiology, Jichi Medical University, 3311-1 Yakushiji, Shimotsuke, Tochigi, 329-0498, Japan; Department of Computational Diagnostic Radiology and Preventive Medicine, The University of Tokyo Hospital, 7-3-1 Hongo, Bunkyo-ku, Tokyo, 113-8655, Japan; Division of Radiology and Biomedical Engineering, Graduate School of Medicine, The University of Tokyo, 7-3-1 Hongo, Bunkyo-ku, Tokyo, 113-8655, Japan; Department of Radiology, Juntendo University School of Medicine, 2-1-1 Hongo, Bunkyo-ku, Tokyo, 113-8421, Japan; Department of Radiology, The University of Tokyo Hospital, 7-3-1 Hongo, Bunkyo-ku, Tokyo, 113-8655, Japan

**Keywords:** Computed tomography, Abdomen, Reference values, Organ volume, Segmentation

## Abstract

**Background:** Large-scale CT-based reference standards for abdominal organ volume, incorporating age, sex, and body size, are limited.

**Purpose:** To establish sex- and age-specific reference distributions for major abdominal organ volumes on non-contrast abdominopelvic CT in a nationwide Japanese cohort to provide a foundation for automated clinical assessment and dose optimization.

**Materials and Methods:** In this retrospective, multicenter study, using the Japan Medical Image Database, we identified all non-contrast abdominopelvic CT examinations performed in 2024. Unique adults with available data on age, sex, height, and weight were included in this study. The final sample comprised 49,764 examinations (26,456 men and 23,308 women) conducted at nine institutions. Automated segmentation (TotalSegmentator v2.10.0) was used to produce organ volumes, excluding hollow viscera. The sex-specific 10th, 25th, 50th, 75th, and 90th percentiles were calculated. Age–volume relationships of body surface area (BSA)-normalized volumes (mL/m^2^) were modeled using natural cubic splines (four degrees of freedom) separately by sex.

**Results:** Median (mL) male vs female volumes were as follows: liver, 1194.7 vs 1024.0; pancreas, 63.6 vs 52.2; spleen, 118.1 vs 95.1; kidneys (total), 268.3 vs 221.2; adrenals (total), 6.6 vs 4.2; iliopsoas (total), 483.4 vs 317.7; prostate, 24.9 (men only). Age–volume relationships of BSA-normalized volumes showed convex patterns for the liver, pancreas, and kidneys in both sexes and for male adrenal glands; lower values in older age groups for the spleen and iliopsoas in both sexes; and higher values in older age groups for the prostate and female adrenal glands.

**Conclusion:** This nationwide Japanese CT cohort provides sex- and age-resolved volumetric reference standards. These standards enable objective identification of abnormalities, support personalized medicine, and facilitate automated AI-based reporting to reduce radiologist workload and optimize radiation dose protocols.

**Key Results:**

- Median volumes (men vs women, mL): liver 1195/1024; pancreas 64/52; spleen 118/95; kidneys 268/221; adrenals 6.6/4.2; iliopsoas 483/318; prostate 25.
- Body surface area–normalized age–volume relationships were convex for liver, pancreas, and kidneys in both sexes and for male adrenal glands.
- Spleen and iliopsoas declined monotonically with age in both sexes, whereas prostate and female adrenal glands increased monotonically.

## Introduction

Advances in computer vision and data science have expanded medical image analysis beyond subjective visual reads to standardized quantitative outputs, including organ volumes, CT attenuation, radiomic descriptors, and AI-derived metrics, yielding clinically useful insights^1,2^. In particular, organ volumetry has been linked to functional assessment, disease course, and prognosis. For example, total kidney volume predicts functional prognosis in autosomal dominant polycystic kidney disease, preoperative liver volume predicts post-hepatectomy outcomes, and pancreatic volume changes are associated with diabetes, underscoring its value as an imaging biomarker^3– 6^. Reference standards describing organ volume distributions in large cohorts are therefore essential as foundational infrastructure for quantitative imaging. Previous CT-based studies, typically involving several hundred to a few thousand individuals, have reported reference values for individual abdominal organs^7–11^. Although recent pioneering works have successfully utilized automated segmentation in relatively large cohorts^10,11^, many studies in this field have historically relied on manual or semi-automated methods^7–9^, which often limit reproducibility and cross-study comparability. Furthermore, nationwide reference data covering multiple abdominal organs and constructed using a single, standardized, and fully automated pipeline remain scarce^12^.

The Japan Medical Image Database (J-MID) is a national cloud-based platform that continuously aggregates routine CT and MR images from 11 major university hospitals in Japan^13^. Leveraging this resource, we extracted non-contrast abdominopelvic CT examinations and applied TotalSegmentator, a publicly available and fully automated deep learning–based segmentation framework, to compute volumes for multiple abdominal organs using a single, consistent pipeline^14^.

Using tens of thousands of examinations, we derived sex- and age-specific organ volume distributions that can serve as nationwide reference standards for clinical interpretation, epidemiologic studies, and downstream AI applications, including automated reporting and radiation dose optimization.

## Methods

This study was conducted following the STROBE (Strengthening the Reporting of Observational Studies in Epidemiology) guidelines^15^.

### Study Design and Data Source

In this retrospective multicenter study, we used data from the J-MID, a nationwide repository that aggregates CT and MRI examinations, together with radiology reports from 11 university hospitals across Japan^13^.

### Patient Selection

The patient selection flowchart used in this study is shown in Figure 1. All abdominopelvic CT examinations performed in 2024 were screened for eligibility. The inclusion criteria were as follows: (1) availability of an axial non-contrast series and (2) coverage of the abdomen and pelvis. Examinations were excluded if they met any of the following conditions: (1) multiple CT examinations obtained from the same patient within 2024 (only the first examination was included); (2) missing demographic or anthropometric information (age, sex, height, or weight) in the DICOM metadata; or (3) out-of-range anthropometric values (age < 20 or > 89 years, height < 1.0 or > 2.0 m, or weight < 30 or > 120 kg). These ranges were also applied to avoid inclusion of outliers or potentially identifiable extreme values when publishing statistical information. After applying these criteria, 49,764 examinations were included in the final analysis.

**Figure 1:**
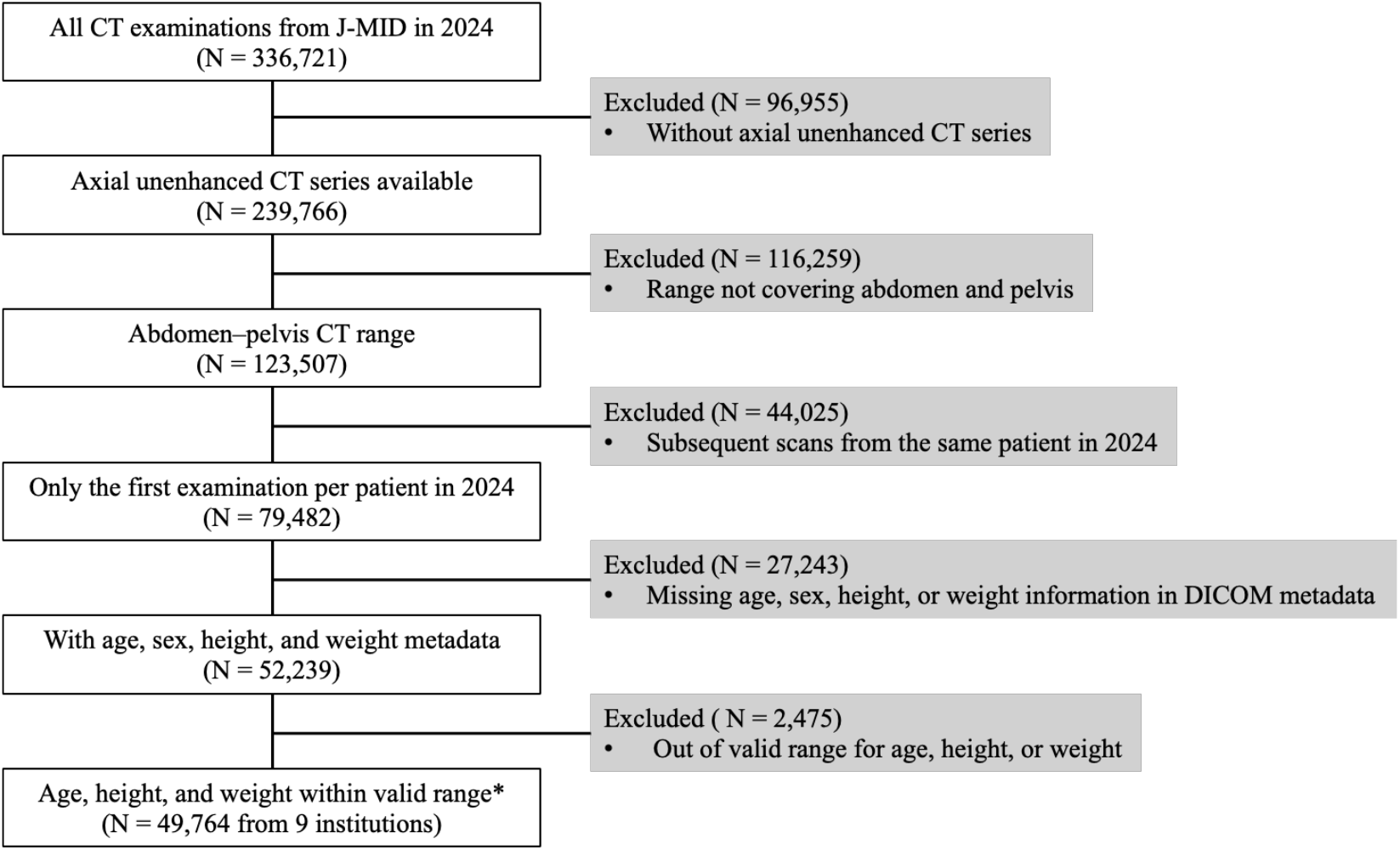
Flowchart of case selection *Valid ranges were defined as age 20–89 years, height 1.0–2.0 m, and weight 30–120 kg.

### CT Acquisition

The included CT examinations were acquired during routine clinical practice at the participating institutions of the J-MID. Scanner models, acquisition parameters, and reconstruction protocols varied among institutions and were not standardized.

### Image Analysis

All the included CT datasets were processed using TotalSegmentator (version 2.10.0) for automated organ segmentation^14^. The processing was performed with the “--fast” option, which resamples the input images to a 3-mm isotropic resolution. The processing pipeline was executed on a Linux workstation equipped with an NVIDIA RTX 6000 Ada GPU under Ubuntu 22.04. Abdominal and pelvic organs were extracted while hollow viscera were excluded from the TotalSegmentator output. The following organs were analyzed: the liver, pancreas, spleen, right and left kidneys, right and left adrenal glands, prostate, and right and left iliopsoas muscles, and the segmented volume was calculated for each organ using custom Python scripts. The performance of TotalSegmentator was validated through a preliminary evaluation using a subset of our cohort (Appendix A).

### Statistical Analysis

Descriptive statistics, including age, height, weight, and body surface area (BSA), were initially calculated to summarize the characteristics of the study cohort by participating institution and sex. The correlations between organ volume and demographic or anthropometric variables (age, sex, height, weight, and BSA) were preliminarily examined, and the results are presented in Appendix B. For each organ, sex-specific organ volumes were calculated as the 10th, 25th, 50th, 75th, and 90th percentiles based on the full study cohort.

In addition, the relationship between age and organ volume normalized based on BSA (mL/m^2^) was visualized separately for males and females. Using all available data, smoothed reference curves were generated using natural cubic spline functions with four degrees of freedom, and the resulting age–volume relationships were visually classified into one of five patterns: higher volumes in older age groups, lower volumes in older age groups, convex (mountain-shaped), concave (valley-shaped), or flat (no apparent change). To safeguard participant privacy and minimize the risk of individual re-identification from uniquely small or large anatomical measurements, the tabulated results and scatter plots focus on the 10th–90th percentile range. This restriction was applied only to data presentation and visualization and did not affect the underlying statistical analyses, which were performed using the full dataset. No disease-specific or regional subgroup analyses were performed, as detailed clinical and sociodemographic information was not uniformly available across institutions.

All statistical analyses and plotting were performed using Python version 3.11.13, with the following packages: statsmodels 0.14.5, pandas 2.3.1, and matplotlib 3.10.5, along with their dependent libraries. Custom scripts used for data preprocessing and summarization are available from the corresponding author upon reasonable request.

### Ethics

This study was approved by the Institutional Review Board of the participating institution (approval number: J24-017). The requirement for written informed consent was waived because of the retrospective nature of the study.

## Results

Overall, 49,764 CT examinations were included in the final analysis (26,456 men and 23,308 women). Of the 11 J-MID participating institutions, nine (A to I) provided eligible cases during the study period, with the number of cases per institution ranging from 171 to 10,491.

The basic demographic and anthropometric characteristics of the study cohort are summarized in Table 1. Information on the CT manufacturer, pixel spacing, and slice thickness is summarized in Table 2. Toshiba scanners accounted for the largest proportion of examinations, followed by Canon, Siemens, GE, and Philips. Although Toshiba Medical Systems has since merged into Canon Medical Systems, manufacturer categories were defined according to the original manufacturer information recorded in the DICOM headers to preserve consistency with acquisition-time metadata.

**Table 1.** Demographic and anthropometric characteristics of the study cohort.

|  | Male | Female |
| --- | --- | --- |
| **Sample Size\*** | 26456<br>A:1311, B:3857, C:110,<br>D:3790, E:4223, F:5536,<br>G:3070, H:2945, I:1614 | 23308<br>A:1322, B:2654, C:61,<br>D:3441, E:3449, F:4955,<br>G:2875, H:3199, I:1352 |
| **Age (year)** | 71 [60, 77] | 67 [54, 76] |
| **Height (m)** | 1.67 [1.63, 1.72] | 1.55 [1.50, 1.59] |
| **Weight (kg)** | 64.9 [57.5, 72.7] | 52.3 [46.1, 60.0] |
| **BSA (m<sup>2</sup>)\*\*** | 1.73 [1.63, 1.84] | 1.50 [1.40, 1.60] |

*All values except sample size are presented as median (25th, 75th percentile).*
*\* A–I indicate anonymized institution identifiers.*
*\*\* Body Surface Area*

**Table 2.** Distribution of CT information.

|  |  |  |
| --- | --- | --- |
| **Manufacturer** | Toshiba | 16504 |
|  | Canon | 14938 |
|  | Siemens | 8730 |
|  | GE | 6849 |
|  | Philips | 2646 |
|  | Others or no Information | 97 |
| **Pixel spacing (mm)** | > 1.0 | 3648 |
|  | 0.5 - 1.0 | 45970 |
|  | < 0.5 | 146 |
| **Slice thickness (mm)** | $\geq 7$ | 15 |
| | $5 \leq , < 7$ | 38814 |
| | $3 \leq , < 5$ | 10743 |
|  | < 3 | 192 |

Sex-specific summaries of organ volumes are presented in Tables 3 and 4.

**Table 3.** Organ volumes in males.

| **Organ** | **p10\*** | **p25\*** | **p50\*** | **p75\*** | **p90\*** |
| --- | --- | --- | --- | --- | --- |
| Liver | 893.3 | 1024.4 | 1194.7 | 1411.4 | 1667.2 |
| Pancreas | 35.7 | 49.1 | 63.6 | 78.5 | 92.4 |
| Spleen | 57.4 | 82.9 | 118.1 | 167.6 | 234.5 |
| Kidney right | 85.5 | 109.7 | 133.3 | 159 | 187.4 |
| Kidney left | 83.1 | 112.7 | 137.9 | 164.4 | 192.3 |
| Kidney total | 177.8 | 221.8 | 268.3 | 317.1 | 369.2 |
| Adrenal gland rt. | 1.5 | 2.2 | 3 | 3.7 | 4.5 |
| Adrenal gland lt. | 1.7 | 2.8 | 3.7 | 4.6 | 5.6 |
| Adrenal gland Total | 3.6 | 5.1 | 6.6 | 8.2 | 9.8 |
| Prostate | 10.9 | 19.4 | 24.9 | 30.3 | 36.3 |
| Iliopsoas muscle rt. | 166.8 | 199.9 | 237.6 | 279.3 | 323.2 |
| Iliopsoas muscle lt. | 170.2 | 205.5 | 246.2 | 290.5 | 334.5 |
| Iliopsoas muscle Total | 339.6 | 406.5 | 483.4 | 568 | 655.1 |

The units of the values are milliliters (mL).
\*p10, p25, p50, p75, and p90 indicate percentiles.

**Table 4.** Organ volumes in females.

| **Organ** | **p10\*** | **p25\*** | **p50\*** | **p75\*** | **p90\*** |
| --- | --- | --- | --- | --- | --- |
| Liver | 776.1 | 886 | 1024 | 1203.9 | 1439.9 |
| Pancreas | 27.9 | 40 | 52.2 | 64.5 | 75.8 |
| Spleen | 48 | 67.4 | 95.1 | 134.1 | 187.3 |
| Kidney right | 72.6 | 90.8 | 108.7 | 128.7 | 151.5 |
| Kidney left | 73.2 | 94.9 | 114.6 | 135.2 | 158.1 |
| Kidney total | 150.7 | 185.5 | 221.2 | 260 | 302.6 |
| Adrenal gland rt. | 0.7 | 1.2 | 1.9 | 2.6 | 3.3 |
| Adrenal gland lt. | 0.6 | 1.5 | 2.4 | 3.2 | 4 |
| Adrenal gland Total | 1.7 | 2.9 | 4.2 | 5.7 | 7.1 |
| Iliopsoas muscle rt. | 112.3 | 133.9 | 156.5 | 179.9 | 202.9 |
| Iliopsoas muscle lt. | 114.6 | 137.9 | 161.6 | 186.3 | 210.8 |
| Iliopsoas muscle Total | 230.1 | 272.3 | 317.7 | 364.6 | 411.9 |

\*p10, p25, p50, p75, and p90 indicate percentiles.

For paired organs such as the kidneys, adrenal glands, and iliopsoas muscles, the total volumes were additionally calculated by summing the values for the right and left sides.

Figure 2 shows the relationship between age and body surface area–normalized organ volume (mL/m^2^) for each organ. Based on visual inspection of the spline curves, the liver, pancreas, and kidneys exhibited similar convex patterns in both sexes, with largely overlapping distributions between males and females. In contrast, sex differences were more pronounced for the adrenal glands and iliopsoas muscle. Adrenal gland volumes showed a convex pattern in males and a monotonic increase in females, whereas iliopsoas volumes were lower in older age groups in both sexes. Prostate volumes in males demonstrated a wider range of values in older age groups, while spleen volumes declined with age and showed minimal separation between sexes.

**Figure 2:**
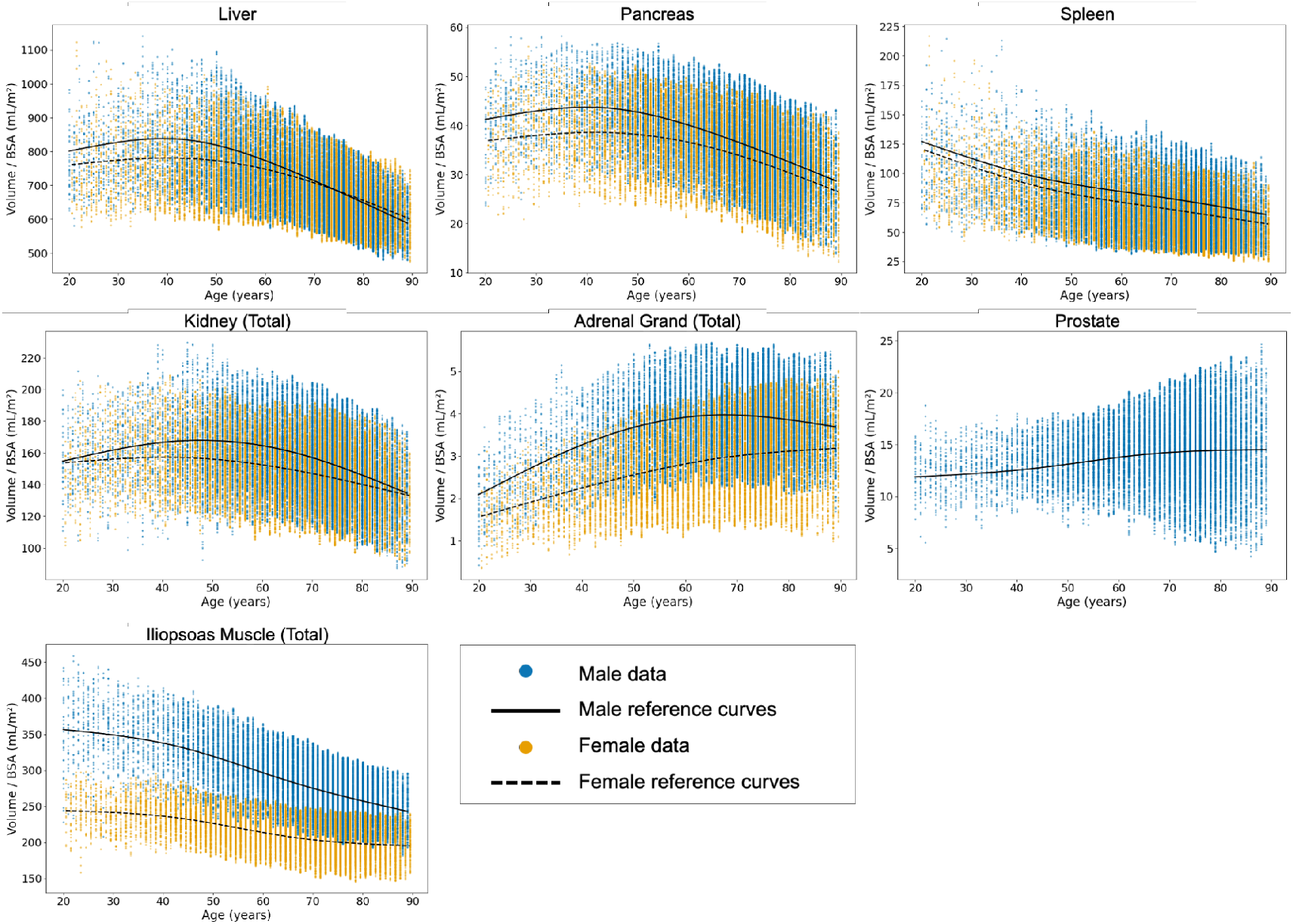
Relationship between age and body surface area–normalized organ volume Scatterplots showing the relationship between age and organ volume normalized based on body surface area in mL/m^2^ for each organ. The blue and orange dots represent male and female data points, respectively. Only data within the 10th–90th percentiles were plotted. The black solid and dashed lines indicate the smoothed reference curves generated using natural cubic spline functions with four degrees of freedom for males and females, respectively.

## Discussion

Using approximately 50,000 routine non-contrast abdominopelvic CT examinations from a nationwide, multi-institutional Japanese platform, this study generated reference ranges and body surface area–normalized plots of organ volume versus age. Distinct cross-sectional age–volume relationships were observed, with convex patterns for the liver, pancreas, and kidneys in both sexes, monotonic declines for the spleen and iliopsoas in both sexes, and sex-specific patterns for the adrenal glands. Prostate volumes in males showed greater variability in older age groups. Appendix C provides summary tables of organ volumes by body surface area and age. Overall, given its sample size and analytical scope across age, sex, and body size, this represents one of the most comprehensive abdominal CT volumetric analyses to date.

Previous CT-based organ volumetry studies have typically involved cohorts of several hundred to a few thousand participants and provided estimates of “normal” organ volumes^7,10,11,15^. For example, Tamura et al. established normative spleen volume references in healthy Japanese adults using automated CT segmentation, and Kim et al. reported population-based and personalized reference intervals for liver and spleen volumes using CT and AI, demonstrating the clinical utility of population-specific reference modeling^10,11^. However, these studies lacked sufficient scale to characterize empirical distributions stratified by age, sex, and body size. Despite the recent availability of large imaging datasets, few—particularly from Western populations—support population-level reference standards integrating these factors^12,16-18^. In this context, the present study represents one approach to addressing this gap through analyses based on nationwide Japanese CT data.

The age–volume relationships observed for the liver, pancreas, and kidneys are consistent with established biological findings, including lower liver size and perfusion in older individuals, reduced kidney cortical volume after body-size adjustment, and peak pancreatic volume in early-to-mid adulthood with lower values in older age groups, particularly among patients with diabetes^8,19,20^. Adrenal volume showed age-related and sex-specific differences consistent with prior studies based on several hundred cases, exhibiting a convex pattern in males and higher values in older age groups in females, underscoring the importance of sex-specific reference curves^9^. Spleen volume was modestly lower in older age groups and reflects body habitus, whereas skeletal muscle metrics, including psoas volume, showed age-related differences with marked sex effects^21,22^. Together, these findings suggest that the cross-sectional reference curves reflect established physiological variation rather than measurement artifacts. Prostate volumes were generally higher in older age groups, with greater variability after approximately age 60, potentially reflecting benign prostatic hyperplasia and volume reduction following treatment^23^.

The establishment of nationwide reference standards has several important implications. Integration of deep learning–based segmentation with these references enables automated organ volume reporting in routine CT, with potential benefits for diagnostic accuracy, inter-observer consistency, and radiologist workload, as highlighted in recent reviews^24^. Beyond routine reporting, age- and body size–adjusted organ volume references may support investigations of imaging-based proxies of biological aging, identification of early or subclinical organ atrophy, and individualized risk stratification and longitudinal monitoring in precision medicine. In addition, organ-specific volumetric data may aid refinement of CT protocols to optimize radiation dose while maintaining diagnostic quality, consistent with Diagnostic Reference Level initiatives^25^. By providing Asian-specific reference values, this study addresses ethnic and body-size differences underrepresented in Western-centric datasets, supporting equity in diagnostic imaging and aging research. Although focused on Japan, these benchmarks may be applicable to other East Asian populations with similar body habitus and provide a basis for international comparisons.

This study has several limitations. First, the cohort comprised individuals undergoing routine clinical imaging rather than healthy volunteers. Although not all abdominal organs are likely abnormal in the same individual, specific pathologies (e.g., cirrhosis or splenomegaly) may have introduced bias and could not be systematically excluded because radiology reports were not uniformly available. Accordingly, these reference standards reflect a routine clinical population rather than a strictly healthy cohort. Second, segmentation errors may have influenced the aggregated results. While TotalSegmentator is a robust framework, non-contrast CT can introduce measurement variability for structures with less clearly defined boundaries. To focus on the central distribution and reduce re-identification risk, tabulated results and visualizations were limited to the 10th–90th percentile range. Variation in slice thickness may also have affected volume estimation, although this reflects real-world clinical practice. Third, disease-specific and regional subgroup analyses were not performed because detailed clinical and sociodemographic data were not consistently available; these factors warrant investigation in future studies. Fourth, the ethnic composition was unbalanced, as foreign residents accounted for approximately 3% of Japan’s population in 2024, and the dataset predominantly represents Japanese individuals^26^. Finally, this cross-sectional study captures population-level differences across age groups in 2024 and does not reflect within-individual longitudinal aging.

In conclusion, using a nationwide Japanese CT dataset, we established sex- and age-specific reference standards for abdominal organ volume and identified age-related differences. These CT-based references provide practical resources for clinical use, epidemiology, and AI development.

## Data Availability

Statistical data from this study will be shared upon reasonable request to the corresponding author. The original clinical data obtained from the J-MID cannot be publicly shared because of ethical and privacy restrictions.

## Abbreviation List

CT: Computed Tomography
AI: Artificial Intelligence
J-MID: Japan Medical Image Database
BSA: Body Surface Area
MR: Magnetic Resonance

## Acknowledgments

We would like to thank the departments of radiology that provided the J-MID database, including Juntendo Univ., Kyushu Univ., Keio Univ., The Univ. of Tokyo, Okayama Univ., Kyoto Univ., Osaka Univ., Tokyo Medical and Dental Univ., Hokkaido Univ., Ehime Univ., and Tokushima Univ.

