## Appendix for "Nationwide Organ Volume Reference Standards and Aging-Related Changes in Abdominal CT from Japan"

### A. Preliminary evaluation of TotalSegmentator

A small-scale preliminary evaluation of automated organ segmentation was conducted using abdominopelvic CT examinations acquired on October 1, 2024. From the available data, 96 examinations were sampled from six institutions, with 16 cases per institution. For each case, segmentation results for the liver, spleen, pancreas, and bilateral kidneys generated by TotalSegmentator were compared with manually created reference labels generated by a single board-certified radiologist, and Dice similarity coefficients were calculated. Institutions in which 16 eligible examinations could not be secured on the same acquisition date were excluded from this preliminary analysis. Overall, the Dice similarity coefficients indicated acceptable segmentation performance across all five organs.

| Organ | Facility* | | | | | |
|---|---|---|---|---|---|---|
| | B | D | E | F | G | H |
| Liver | 0.970 | 0.974 | 0.976 | 0.977 | 0.979 | 0.971 |
| Spleen | 0.944 | 0.952 | 0.958 | 0.964 | 0.967 | 0.958 |
| Kidney Rt. | 0.944 | 0.956 | 0.950 | 0.960 | 0.956 | 0.951 |
| Kidney Lt. | 0.953 | 0.957 | 0.953 | 0.961 | 0.961 | 0.955 |
| Pancreas | 0.854 | 0.885 | 0.891 | 0.890 | 0.901 | 0.887 |

### B. Correlation coefficients with organ volumes

*The row corresponding to BSA is shaded in gray. Red indicates the variable with the highest absolute correlation coefficient with organ volume, and green indicates those within the top three. For all organs analyzed, the absolute correlation coefficient for BSA ranked among the top three.

| | Liver | Pancreas | Spleen | Kidney | Adrenal grand | Prostate | Iliopsoas mucle |
|---|---|---|---|---|---|---|---|
| **Age** | -0.322 | -0.308 | -0.198 | -0.203 | 0.139 | 0.036 | -0.258 |
| **Age^2** | -0.340 | -0.326 | -0.196 | -0.220 | 0.119 | 0.026 | -0.263 |
| **Age^3** | -0.348 | -0.334 | -0.191 | -0.230 | 0.100 | 0.017 | -0.263 |
| **Height** | 0.394 | 0.391 | 0.218 | 0.412 | 0.366 | 0.143 | 0.695 |
| **Height^2** | 0.395 | 0.392 | 0.219 | 0.413 | 0.367 | 0.142 | **0.698** |
| **Height^3** | 0.396 | 0.392 | 0.220 | 0.414 | 0.367 | 0.141 | **0.700** |
| **Weight** | **0.562** | **0.568** | **0.306** | **0.571** | **0.614** | **0.205** | 0.674 |
| **Weight^2** | **0.562** | **0.548** | **0.308** | **0.560** | **0.591** | **0.192** | 0.658 |
| **Weight^3** | 0.548 | 0.518 | 0.304 | 0.536 | 0.554 | 0.176 | 0.625 |
| **BSA** | **0.560** | **0.572** | **0.306** | **0.578** | **0.599** | **0.215** | **0.757** |
| **Sex (M)** | 0.233 | 0.271 | 0.148 | 0.297 | 0.448 | --- | 0.634 |

### C. Summary tables by age and body surface area

*The table presents median values and the 25th and 75th percentiles, with age on the vertical axis and body surface area (BSA) on the horizontal axis.

### Liver

- **Male**

| | 1.3-1.4 | 1.4-1.5 | 1.5-1.6 | 1.6-1.7 | 1.7-1.8 | 1.8-1.9 | 1.9-2.0 | 2.0-2.1 | 2.1-2.2 | 2.2-2.3 |
|---|---|---|---|---|---|---|---|---|---|---|
| **20-24** | | 1109.0 [951.9, 1274.8] | 1151.2 [1040.7, 1308.8] | 1242.6 [1143.2, 1405.1] | 1294.9 [1190.5, 1416.8] | 1453.1 [1322.9, 1561.9] | 1609.0 [1490.2, 1748.3] | | | |
| **25-29** | | | 1148.1 [965.6, 1277.3] | 1224.5 [1108.2, 1363.5] | 1347.6 [1261.5, 1505.1] | 1517.9 [1359.7, 1628.9] | 1607.1 [1314.8, 1726.1] | 1669.2 [1438.6, 1861.1] | | |
| **30-34** | | | 1168.6 [1069.7, 1280.9] | 1414.7 [1230.9, 1521.4] | 1385.9 [1242.1, 1518.5] | 1411.9 [1249.3, 1638.8] | 1561.5 [1397.7, 1806.6] | 1686.5 [1494.4, 1896.2] | 1839.9 [1726.6, 2066.0] | |
| **35-39** | | | 1261.7 [1069.7, 1388.3] | 1317.6 [1133.0, 1521.6] | 1336.6 [1223.5, 1522.5] | 1510.7 [1396.3, 1656.8] | 1556.5 [1401.7, 1758.9] | 1667.2 [1544.2, 1954.9] | 1954.6 [1830.5, 2230.7] | |
| **40-44** | | 1147.8 [1031.0, 1243.5] | 1169.0 [1058.0, 1376.7] | 1219.0 [1121.7, 1413.0] | 1329.0 [1207.0, 1496.1] | 1477.6 [1331.0, 1724.4] | 1622.3 [1427.1, 1833.1] | 1769.0 [1514.0, 1955.9] | 1888.4 [1603.2, 2167.0] | |
| **45-49** | | 1040.5 [934.9, 1174.0] | 1139.8 [1056.3, 1230.1] | 1247.7 [1113.4, 1448.8] | 1354.8 [1217.6, 1513.8] | 1414.0 [1285.6, 1652.5] | 1578.9 [1385.6, 1801.9] | 1714.5 [1467.9, 1918.8] | 1805.0 [1676.4, 2220.8] | 2017.0 [1805.2, 2301.6] |
| **50-54** | | 1032.9 [838.8, 1155.7] | 1176.4 [1021.4, 1334.6] | 1278.6 [1140.3, 1425.2] | 1335.0 [1190.5, 1553.2] | 1434.1 [1265.2, 1644.8] | 1592.4 [1388.8, 1778.2] | 1767.6 [1576.9, 1982.0] | 1925.6 [1697.1, 2163.5] | 1950.6 [1718.5, 2319.7] |
| **55-59** | | 1001.1 [859.5, 1100.0] | 1125.3 [998.7, 1296.0] | 1206.3 [1101.4, 1373.5] | 1347.4 [1199.4, 1498.3] | 1404.9 [1263.2, 1564.9] | 1517.4 [1357.2, 1766.8] | 1638.7 [1489.6, 1857.2] | 1691.0 [1468.1, 1965.6] | 1853.2 [1657.2, 2004.3] |
| **60-64** | 896.0 [836.4, 930.7] | 1025.2 [923.2, 1129.7] | 1068.4 [983.9, 1215.4] | 1176.8 [1066.1, 1325.4] | 1306.2 [1143.0, 1455.9] | 1394.6 [1249.9, 1573.8] | 1489.0 [1321.4, 1672.0] | 1552.6 [1369.4, 1778.3] | 1691.5 [1471.5, 1956.4] | 1852.0 [1641.7, 2488.5] |
| **65-69** | 938.9 [839.5, 1045.3] | 993.5 [881.9, 1112.1] | 1078.1 [961.5, 1198.9] | 1143.8 [1041.1, 1261.0] | 1240.1 [1118.7, 1407.2] | 1323.4 [1194.0, 1494.3] | 1440.0 [1281.0, 1625.9] | 1529.1 [1367.6, 1707.0] | 1665.3 [1408.8, 1965.7] | 1831.5 [1724.6, 2002.5] |
| **70-74** | 857.9 [751.6, 996.2] | 972.5 [858.5, 1081.4] | 1034.5 [934.1, 1159.4] | 1109.7 [997.3, 1242.5] | 1196.0 [1068.6, 1345.3] | 1287.4 [1162.8, 1435.5] | 1394.0 [1238.3, 1568.6] | 1472.2 [1294.1, 1689.3] | 1482.8 [1305.0, 1763.1] | |
| **75-79** | 866.1 [748.4, 960.1] | 923.5 [831.8, 1043.6] | 995.2 [885.9, 1116.5] | 1072.8 [958.5, 1200.3] | 1142.9 [1023.8, 1285.2] | 1229.6 [1112.9, 1398.3] | 1282.5 [1169.6, 1452.4] | 1411.3 [1270.3, 1567.6] | 1541.9 [1440.5, 1656.2] | |
| **80-84** | 797.9 [695.8, 927.6] | 891.4 [796.5, 1000.2] | 959.9 [862.0, 1060.6] | 1028.5 [918.6, 1146.1] | 1102.2 [995.3, 1227.4] | 1144.3 [1036.0, 1264.0] | 1230.5 [1103.8, 1417.6] | 1301.8 [1197.8, 1567.1] | | |
| **85-89** | 821.3 [714.1, 886.2] | 881.7 [775.4, 957.3] | 932.4 [836.5, 1021.7] | 990.5 [894.4, 1097.6] | 1071.8 [954.2, 1188.3] | 1128.6 [1029.1, 1263.0] | 1237.7 [1073.7, 1378.5] | | | |

- **Female**

| | 1.1-1.2 | 1.2-1.3 | 1.3-1.4 | 1.4-1.5 | 1.5-1.6 | 1.6-1.7 | 1.7-1.8 | 1.8-1.9 | 1.9-2.0 | 2.0-2.1 |
|---|---|---|---|---|---|---|---|---|---|---|
| **20-24** | | 904.2 [812.7, 939.6] | 1024.3 [949.1, 1140.6] | 1025.2 [927.5, 1147.0] | 1122.8 [1010.6, 1279.8] | 1184.6 [1046.8, 1449.4] | 1253.3 [1156.5, 1446.2] | | | |
| **25-29** | | 925.2 [812.4, 1011.2] | 989.0 [904.5, 1064.3] | 1061.4 [940.4, 1172.2] | 1097.3 [999.1, 1274.8] | 1283.6 [1084.4, 1427.0] | 1513.3 [1204.8, 1893.9] | 1677.3 [1371.1, 1767.6] | | |
| **30-34** | | 909.8 [840.2, 1032.6] | 974.8 [878.1, 1147.8] | 1053.4 [971.6, 1180.9] | 1143.1 [1020.9, 1288.8] | 1275.9 [1134.9, 1487.0] | 1332.1 [1234.9, 1495.4] | 1413.5 [1237.8, 1661.9] | 2094.9 [1946.6, 2220.2] | |
| **35-39** | | 962.3 [880.2, 1019.8] | 944.2 [892.9, 1052.1] | 1049.4 [960.5, 1171.8] | 1104.6 [986.3, 1261.4] | 1259.3 [1136.1, 1357.5] | 1304.4 [1182.6, 1518.6] | 1477.9 [1305.7, 1655.9] | 1592.5 [1509.1, 1983.1] | |
| **40-44** | | 865.1 [773.7, 991.1] | 987.3 [860.5, 1098.8] | 1039.1 [929.4, 1149.5] | 1118.2 [1024.4, 1252.0] | 1230.3 [1105.2, 1350.9] | 1372.0 [1224.3, 1509.7] | 1668.9 [1438.1, 1841.1] | 1546.5 [1433.4, 1793.0] | 1953.3 [1725.3, 2177.2] |
| **45-49** | | 888.2 [773.4, 1052.5] | 975.1 [863.4, 1083.6] | 1027.1 [930.0, 1154.5] | 1125.6 [1014.9, 1266.6] | 1222.2 [1088.2, 1404.8] | 1324.0 [1174.9, 1506.7] | 1570.1 [1325.0, 1755.1] | 1659.9 [1526.7, 1925.6] | 1941.8 [1571.6, 2068.7] |
| **50-54** | | 900.8 [808.4, 1012.1] | 969.3 [864.0, 1062.9] | 1026.0 [924.8, 1167.6] | 1097.5 [995.1, 1217.1] | 1233.8 [1082.4, 1416.5] | 1367.8 [1192.9, 1602.4] | 1537.3 [1353.5, 1751.2] | 1786.2 [1503.9, 2052.2] | 1827.8 [1488.1, 2018.1] |
| **55-59** | | 891.0 [783.4, 963.0] | 952.7 [851.7, 1061.6] | 1005.6 [902.4, 1137.4] | 1099.3 [985.9, 1239.0] | 1200.0 [1050.4, 1355.3] | 1370.7 [1218.5, 1556.3] | 1538.9 [1335.6, 1720.3] | 1647.8 [1481.3, 1778.0] | |
| **60-64** | 860.4 [738.4, 956.1] | 872.0 [799.7, 986.9] | 937.3 [833.6, 1052.1] | 1016.3 [911.6, 1125.3] | 1080.4 [971.9, 1227.8] | 1206.3 [1071.9, 1344.3] | 1347.0 [1175.3, 1577.0] | 1407.8 [1242.1, 1619.3] | 1592.5 [1488.7, 1777.2] | 1640.3 [1418.2, 1877.7] |
| **65-69** | 791.0 [713.8, 879.3] | 862.6 [771.8, 980.5] | 918.5 [833.4, 1029.6] | 980.4 [882.7, 1109.8] | 1074.4 [964.0, 1205.4] | 1177.7 [1047.0, 1340.0] | 1247.7 [1116.6, 1451.8] | 1407.5 [1224.7, 1645.0] | 1501.6 [1358.7, 1655.3] | |
| **70-74** | 791.4 [681.3, 938.6] | 824.8 [734.7, 907.9] | 886.8 [798.8, 991.2] | 955.0 [860.4, 1063.7] | 1019.3 [914.7, 1170.6] | 1139.1 [1030.5, 1283.7] | 1220.0 [1084.5, 1388.5] | 1317.8 [1171.9, 1551.2] | 1478.0 [1260.2, 1652.5] | |
| **75-79** | 769.9 [667.6, 858.6] | 814.1 [720.7, 909.3] | 866.7 [782.8, 965.4] | 929.8 [831.3, 1036.3] | 996.1 [903.3, 1122.5] | 1112.4 [1001.6, 1262.4] | 1161.2 [1031.1, 1334.9] | 1183.9 [1048.2, 1508.9] | | |
| **80-84** | 697.5 [636.9, 766.3] | 797.2 [706.3, 903.0] | 842.4 [760.8, 947.5] | 907.6 [809.3, 1013.6] | 989.7 [875.4, 1101.1] | 1076.3 [970.6, 1171.7] | 1191.4 [1028.0, 1264.0] | | | |
| **85-89** | 700.1 [619.5, 825.5] | 764.1 [696.5, 852.9] | 829.2 [720.1, 920.6] | 885.5 [797.0, 980.3] | 981.9 [839.2, 1078.2] | 992.7 [904.9, 1188.8] | 957.8 [919.8, 1314.4] | | | |

### Pancreas

- **Male**

| | 1.3-1.4 | 1.4-1.5 | 1.5-1.6 | 1.6-1.7 | 1.7-1.8 | 1.8-1.9 | 1.9-2.0 | 2.0-2.1 | 2.1-2.2 | 2.2-2.3 |
|---|---|---|---|---|---|---|---|---|---|---|
| **20-24** | | 55.7 [48.3, 65.6] | 52.8 [45.9, 66.3] | 68.3 [61.0, 76.7] | 70.4 [67.8, 80.7] | 77.4 [72.3, 82.2] | 85.4 [83.1, 93.0] | | | |
| **25-29** | | | 56.9 [50.2, 72.4] | 65.6 [58.5, 76.4] | 75.1 [66.0, 85.5] | 85.7 [71.6, 97.1] | 86.2 [80.2, 98.1] | 89.5 [77.7, 95.3] | | |
| **30-34** | | | 68.6 [57.2, 79.6] | 70.1 [58.0, 82.7] | 74.4 [63.9, 84.6] | 82.8 [68.4, 93.7] | 91.5 [81.5, 105.5] | 96.7 [85.3, 103.0] | 100.4 [77.5, 115.1] | |
| **35-39** | | | 64.2 [47.6, 78.1] | 72.5 [60.7, 84.9] | 79.9 [68.2, 90.1] | 82.2 [73.5, 93.3] | 89.8 [79.8, 97.1] | 92.7 [81.3, 106.4] | 109.6 [98.2, 120.0] | |
| **40-44** | | 67.0 [49.2, 71.3] | 58.8 [48.7, 75.3] | 68.5 [61.9, 79.9] | 75.6 [64.8, 87.5] | 82.4 [71.4, 91.5] | 87.3 [77.9, 97.1] | 88.7 [72.0, 97.7] | 96.3 [85.3, 110.3] | |
| **45-49** | | 46.8 [35.0, 64.8] | 63.4 [46.7, 76.4] | 66.9 [57.6, 77.1] | 79.5 [69.9, 89.2] | 80.6 [68.3, 92.6] | 86.3 [75.8, 99.3] | 91.9 [79.6, 101.6] | 103.1 [87.9, 112.0] | 95.3 [79.0, 114.6] |
| **50-54** | | 49.4 [34.5, 66.2] | 58.7 [48.2, 70.5] | 68.1 [56.2, 79.7] | 73.6 [61.8, 87.4] | 79.4 [67.5, 90.4] | 85.8 [73.5, 97.9] | 92.3 [76.0, 106.4] | 100.5 [88.5, 112.6] | 100.1 [87.2, 112.6] |
| **55-59** | | 46.2 [34.4, 58.1] | 57.4 [45.9, 65.9] | 64.0 [51.3, 74.2] | 72.0 [60.9, 84.2] | 78.3 [65.1, 90.3] | 81.1 [67.2, 92.9] | 88.2 [73.1, 101.5] | 92.1 [79.2, 101.4] | 89.7 [82.4, 99.1] |
| **60-64** | 41.4 [29.1, 48.2] | 43.4 [30.2, 53.5] | 53.8 [41.0, 67.6] | 63.5 [51.5, 74.8] | 70.1 [58.6, 82.5] | 75.5 [63.4, 86.8] | 79.3 [66.8, 93.0] | 82.2 [71.7, 96.9] | 94.6 [77.0, 104.8] | 103.7 [85.5, 123.6] |
| **65-69** | 33.8 [25.9, 45.0] | 44.8 [33.7, 58.1] | 52.8 [41.3, 66.3] | 59.8 [48.8, 71.3] | 67.0 [55.5, 79.5] | 71.7 [60.2, 83.9] | 77.2 [63.3, 89.1] | 81.8 [66.7, 94.5] | 81.2 [70.5, 89.8] | 89.1 [67.0, 104.4] |
| **70-74** | 37.1 [23.5, 47.1] | 43.8 [28.4, 57.5] | 52.8 [40.6, 65.1] | 58.6 [46.8, 70.7] | 64.9 [53.0, 75.9] | 69.7 [57.9, 81.7] | 73.4 [61.3, 85.1] | 76.3 [65.3, 88.8] | 75.5 [62.3, 95.7] | |
| **75-79** | 30.5 [23.8, 42.0] | 43.2 [32.4, 54.8] | 49.1 [38.2, 61.0] | 54.9 [44.7, 66.6] | 61.0 [49.2, 72.5] | 65.1 [52.8, 76.4] | 69.4 [57.6, 79.1] | 74.5 [62.2, 84.4] | 82.5 [62.4, 90.4] | |
| **80-84** | 27.9 [19.0, 46.9] | 41.0 [29.2, 52.0] | 47.3 [35.7, 59.3] | 54.1 [42.7, 65.2] | 57.4 [47.0, 69.8] | 59.1 [50.0, 69.7] | 64.1 [51.0, 79.2] | 72.8 [57.6, 88.6] | | |
| **85-89** | 36.4 [20.9, 50.3] | 37.4 [26.8, 50.0] | 47.6 [34.7, 57.7] | 48.8 [37.7, 58.8] | 55.5 [44.9, 65.6] | 56.8 [44.7, 71.2] | 69.0 [44.4, 77.0] | | | |

- **Female**

| | 1.1-1.2 | 1.2-1.3 | 1.3-1.4 | 1.4-1.5 | 1.5-1.6 | 1.6-1.7 | 1.7-1.8 | 1.8-1.9 | 1.9-2.0 | 2.0-2.1 |
|---|---|---|---|---|---|---|---|---|---|---|
| **20-24** | | 37.7 [10.6, 51.5] | 48.1 [40.6, 59.8] | 49.3 [43.9, 61.3] | 57.7 [49.3, 67.1] | 62.3 [51.8, 72.3] | 65.3 [51.3, 73.6] | | | |
| **25-29** | | 43.4 [29.3, 51.9] | 47.8 [39.5, 61.3] | 55.6 [44.1, 61.3] | 61.2 [52.5, 73.7] | 66.8 [61.1, 72.6] | 70.7 [67.3, 82.8] | 79.1 [66.0, 87.2] | | |
| **30-34** | | 44.0 [22.3, 50.9] | 49.0 [39.5, 58.0] | 54.4 [44.5, 62.4] | 59.2 [49.4, 70.3] | 69.0 [61.7, 80.0] | 72.3 [63.8, 86.6] | 73.6 [65.2, 76.4] | 98.0 [93.2, 107.4] | |
| **35-39** | | 43.0 [28.4, 53.6] | 47.7 [40.2, 54.3] | 54.4 [47.3, 63.3] | 63.0 [53.5, 72.5] | 66.6 [56.1, 74.8] | 70.8 [62.6, 75.2] | 80.2 [71.9, 87.6] | 76.4 [68.7, 83.1] | |
| **40-44** | | 34.2 [20.1, 44.9] | 45.1 [33.4, 54.3] | 51.5 [43.1, 61.4] | 60.9 [52.4, 70.9] | 66.4 [58.9, 77.4] | 71.4 [61.5, 81.7] | 83.0 [69.1, 91.2] | 83.8 [71.9, 93.4] | 88.1 [76.6, 103.4] |
| **45-49** | | 33.1 [24.5, 40.7] | 46.2 [34.9, 56.5] | 52.8 [40.2, 62.7] | 59.2 [50.5, 69.3] | 68.6 [57.0, 78.2] | 74.7 [65.9, 83.7] | 77.0 [69.1, 88.3] | 79.9 [65.3, 88.6] | 84.1 [75.3, 92.8] |
| **50-54** | | 39.5 [25.1, 48.3] | 43.7 [30.3, 55.5] | 53.1 [42.0, 62.3] | 60.0 [50.7, 69.5] | 65.2 [55.7, 74.3] | 71.5 [63.6, 80.6] | 76.0 [65.5, 88.1] | 78.5 [69.6, 88.9] | 75.0 [60.2, 88.2] |
| **55-59** | | 38.0 [29.5, 45.3] | 44.5 [32.9, 52.5] | 50.6 [41.0, 61.4] | 59.9 [48.8, 69.0] | 64.5 [53.7, 74.9] | 68.8 [57.1, 77.6] | 75.8 [61.7, 84.3] | 73.9 [66.7, 87.3] | |
| **60-64** | 32.7 [16.0, 36.6] | 34.2 [22.8, 48.2] | 44.6 [34.8, 54.3] | 52.8 [42.7, 62.3] | 57.6 [48.0, 66.5] | 63.3 [54.4, 74.0] | 66.9 [56.1, 76.0] | 69.0 [55.8, 77.8] | 74.7 [65.4, 81.8] | 71.5 [61.9, 94.3] |
| **65-69** | 33.3 [24.2, 46.0] | 36.2 [23.2, 47.1] | 44.9 [34.2, 54.5] | 51.4 [41.3, 60.3] | 56.8 [47.9, 65.3] | 61.3 [51.2, 69.6] | 63.9 [50.1, 72.3] | 65.5 [53.2, 77.7] | 69.6 [62.1, 82.2] | |
| **70-74** | 31.9 [23.8, 46.0] | 35.1 [22.3, 46.0] | 41.7 [32.6, 51.7] | 48.7 [38.8, 57.5] | 54.2 [44.5, 63.7] | 57.6 [46.9, 70.1] | 58.2 [50.2, 69.9] | 64.2 [48.5, 74.5] | 58.8 [52.1, 69.7] | |
| **75-79** | 28.5 [17.7, 42.2] | 31.8 [20.4, 45.6] | 40.1 [30.1, 50.1] | 46.5 [36.9, 55.4] | 50.5 [41.1, 60.9] | 55.9 [44.9, 65.1] | 56.2 [44.2, 63.5] | 55.2 [47.9, 65.5] | | |
| **80-84** | 29.7 [19.5, 38.9] | 32.0 [20.2, 42.3] | 39.4 [29.3, 49.6] | 44.0 [35.2, 53.4] | 48.1 [38.4, 56.6] | 51.3 [41.9, 60.4] | 55.3 [42.7, 63.8] | | | |
| **85-89** | 29.8 [19.1, 39.6] | 31.7 [21.2, 42.3] | 36.5 [27.2, 46.1] | 42.2 [32.6, 51.0] | 45.4 [35.8, 57.2] | 46.6 [35.6, 53.9] | 48.6 [29.8, 59.8] | | | |

### Spleen

- **Male**

| | 1.3-1.4 | 1.4-1.5 | 1.5-1.6 | 1.6-1.7 | 1.7-1.8 | 1.8-1.9 | 1.9-2.0 | 2.0-2.1 | 2.1-2.2 | 2.2-2.3 |
|---|---|---|---|---|---|---|---|---|---|---|
| **20-24** | | 178.8 [103.3, 234.3] | 161.1 [104.3, 210.8] | 165.1 [142.2, 257.5] | 181.8 [149.6, 220.2] | 198.3 [146.8, 227.8] | 247.9 [220.5, 264.2] | | | |
| **25-29** | | | 125.3 [96.2, 179.8] | 145.4 [118.4, 221.0] | 161.7 [112.6, 201.8] | 193.3 [154.0, 253.9] | 164.7 [145.4, 279.0] | 241.7 [195.0, 291.3] | | |
| **30-34** | | | 155.5 [111.0, 190.0] | 158.1 [128.7, 219.2] | 151.6 [120.9, 199.3] | 166.8 [129.2, 209.7] | 197.3 [146.8, 232.3] | 212.8 [159.9, 278.9] | 206.7 [167.2, 276.1] | |
| **35-39** | | | 134.7 [97.6, 193.3] | 139.6 [104.8, 241.7] | 153.1 [105.8, 221.2] | 166.5 [118.4, 207.6] | 180.3 [142.7, 222.0] | 192.7 [160.6, 232.5] | 285.5 [193.1, 407.0] | |
| **40-44** | | 137.1 [93.8, 205.3] | 118.7 [83.7, 201.2] | 130.1 [87.4, 176.9] | 144.9 [108.0, 192.2] | 150.7 [114.3, 199.7] | 177.0 [126.1, 228.5] | 197.6 [144.9, 263.7] | 179.7 [148.6, 235.2] | |
| **45-49** | | 89.2 [63.3, 144.7] | 98.8 [70.0, 139.6] | 114.3 [87.0, 167.0] | 131.6 [99.3, 187.5] | 148.8 [112.4, 196.9] | 150.4 [119.1, 193.1] | 168.3 [131.0, 227.8] | 236.2 [183.2, 330.4] | 239.4 [169.2, 375.0] |
| **50-54** | | 90.8 [66.5, 151.6] | 102.3 [70.3, 140.2] | 107.9 [79.4, 152.6] | 123.9 [91.0, 168.3] | 147.1 [104.9, 182.2] | 154.3 [118.1, 206.9] | 192.2 [144.3, 244.5] | 208.1 [160.0, 251.8] | 210.5 [190.1, 277.1] |
| **55-59** | | 94.4 [65.4, 137.6] | 90.2 [58.0, 138.8] | 105.1 [76.4, 147.1] | 120.0 [89.2, 162.1] | 131.9 [95.9, 175.0] | 147.5 [112.0, 197.7] | 171.2 [124.1, 232.1] | 169.1 [121.4, 252.0] | 233.1 [173.3, 274.7] |
| **60-64** | 79.4 [63.5, 131.3] | 92.2 [53.3, 114.2] | 90.4 [63.6, 133.0] | 96.7 [70.0, 135.7] | 116.5 [86.8, 164.2] | 135.6 [101.1, 179.3] | 148.9 [111.5, 203.4] | 163.0 [123.7, 226.7] | 189.0 [141.6, 229.0] | 200.3 [145.0, 252.1] |
| **65-69** | 77.7 [48.8, 119.5] | 84.2 [51.9, 112.1] | 89.9 [62.2, 144.3] | 101.6 [75.6, 144.2] | 121.9 [87.0, 170.1] | 132.0 [96.1, 183.0] | 145.6 [106.8, 200.1] | 158.6 [115.6, 222.9] | 170.7 [108.0, 215.1] | 214.3 [193.2, 252.9] |
| **70-74** | 73.5 [49.6, 103.7] | 84.0 [55.3, 121.2] | 91.3 [66.7, 128.5] | 103.6 [71.9, 139.2] | 116.4 [84.5, 159.2] | 130.6 [94.8, 181.3] | 146.2 [105.6, 194.4] | 171.4 [127.2, 228.8] | 187.3 [148.1, 296.1] | |
| **75-79** | 77.2 [44.2, 106.1] | 85.9 [57.4, 124.2] | 92.1 [67.0, 129.8] | 107.6 [77.8, 146.5] | 116.6 [84.9, 159.6] | 125.7 [94.6, 172.4] | 125.9 [94.9, 168.1] | 147.6 [117.4, 210.9] | 187.4 [146.4, 221.1] | |
| **80-84** | 67.4 [49.4, 97.6] | 82.3 [53.1, 113.5] | 92.6 [66.5, 131.4] | 102.0 [74.0, 141.7] | 114.3 [84.4, 151.0] | 115.7 [90.5, 155.5] | 136.8 [105.7, 186.2] | 169.5 [120.9, 218.0] | | |
| **85-89** | 60.8 [48.2, 90.8] | 80.1 [55.5, 117.2] | 90.3 [62.9, 122.5] | 95.5 [69.4, 132.4] | 106.6 [77.4, 136.3] | 119.5 [93.0, 164.7] | 133.4 [95.8, 177.7] | | | |

- **Female**

| | 1.1-1.2 | 1.2-1.3 | 1.3-1.4 | 1.4-1.5 | 1.5-1.6 | 1.6-1.7 | 1.7-1.8 | 1.8-1.9 | 1.9-2.0 | 2.0-2.1 |
|---|---|---|---|---|---|---|---|---|---|---|
| **20-24** | | 147.4 [94.3, 185.8] | 121.7 [93.1, 175.9] | 133.7 [100.9, 164.9] | 144.9 [120.0, 185.6] | 174.1 [136.3, 184.6] | 147.9 [142.2, 191.0] | | | |
| **25-29** | | 123.1 [77.5, 167.2] | 117.0 [90.0, 142.0] | 126.2 [94.4, 161.9] | 147.6 [115.4, 186.1] | 151.1 [124.6, 185.1] | 191.1 [144.2, 283.3] | 197.7 [170.0, 216.1] | | |
| **30-34** | | 90.8 [66.5, 116.9] | 120.2 [95.2, 170.4] | 126.3 [94.1, 167.3] | 136.9 [115.0, 192.6] | 147.2 [113.5, 200.4] | 161.9 [137.1, 195.4] | 152.6 [116.3, 207.5] | 246.7 [219.6, 285.3] | |
| **35-39** | | 77.2 [56.0, 99.7] | 99.1 [80.7, 129.8] | 117.2 [94.3, 144.8] | 131.1 [101.3, 171.6] | 140.7 [109.7, 186.4] | 153.6 [121.1, 181.7] | 184.3 [145.6, 222.8] | 206.6 [171.2, 264.2] | |
| **40-44** | | 76.7 [69.2, 104.2] | 99.7 [74.2, 140.0] | 104.0 [83.5, 143.6] | 117.4 [91.4, 149.7] | 136.0 [106.1, 173.4] | 153.8 [123.3, 189.0] | 172.3 [134.3, 221.9] | 148.8 [121.7, 216.9] | 209.9 [178.8, 309.0] |
| **45-49** | | 84.5 [59.7, 143.2] | 92.5 [72.2, 126.6] | 100.4 [76.7, 130.4] | 109.2 [89.2, 143.0] | 124.6 [96.8, 167.6] | 135.0 [106.0, 172.2] | 164.4 [123.0, 181.6] | 195.6 [140.5, 230.4] | 199.0 [186.0, 273.7] |
| **50-54** | | 82.6 [60.0, 122.4] | 79.8 [56.9, 112.4] | 93.9 [72.2, 127.9] | 108.8 [86.8, 137.6] | 121.1 [95.0, 156.4] | 136.9 [102.4, 190.5] | 172.2 [132.6, 229.7] | 193.6 [157.0, 230.1] | 159.4 [152.4, 221.1] |
| **55-59** | | 70.5 [46.9, 101.3] | 80.9 [60.5, 114.8] | 91.3 [67.6, 120.8] | 102.6 [75.5, 140.0] | 109.6 [84.6, 144.2] | 119.3 [87.9, 163.3] | 160.9 [121.5, 207.3] | 172.9 [126.9, 205.5] | |
| **60-64** | 84.1 [51.5, 127.8] | 82.7 [55.7, 121.7] | 82.0 [53.6, 104.0] | 85.3 [62.0, 120.4] | 96.7 [71.7, 131.3] | 109.4 [80.6, 151.9] | 129.2 [94.3, 166.8] | 140.1 [105.0, 179.2] | 179.0 [146.6, 217.5] | 202.0 [146.1, 252.6] |
| **65-69** | 80.9 [36.1, 117.1] | 75.6 [56.8, 103.8] | 79.9 [60.7, 109.4] | 85.5 [63.9, 117.7] | 94.4 [70.9, 124.3] | 108.0 [80.1, 144.2] | 120.9 [82.6, 159.6] | 137.0 [106.7, 174.9] | 142.3 [112.9, 166.7] | |
| **70-74** | 68.0 [51.5, 99.0] | 63.4 [45.6, 93.3] | 74.2 [52.7, 101.5] | 81.3 [61.2, 109.4] | 93.2 [69.1, 125.7] | 108.6 [81.3, 142.5] | 114.7 [86.0, 153.7] | 130.1 [97.1, 159.3] | 135.7 [110.3, 182.2] | |
| **75-79** | 62.0 [34.4, 93.0] | 63.6 [43.1, 92.0] | 72.5 [52.2, 100.1] | 80.5 [60.1, 109.7] | 89.6 [65.5, 118.5] | 99.7 [74.8, 139.0] | 110.9 [83.0, 136.4] | 155.0 [117.6, 203.9] | | |
| **80-84** | 56.7 [41.6, 68.9] | 63.2 [47.5, 95.2] | 69.6 [50.9, 98.1] | 76.8 [56.9, 106.0] | 85.7 [64.2, 118.1] | 85.8 [61.8, 123.1] | 113.8 [89.8, 143.5] | | | |
| **85-89** | 57.8 [35.8, 83.7] | 58.4 [44.1, 78.7] | 71.7 [51.8, 97.3] | 74.9 [55.1, 99.8] | 83.8 [61.5, 110.6] | 91.2 [69.1, 123.7] | 118.6 [90.8, 143.7] | | | |

### Kidney (Rt.)

- **Male**

| | 1.3-1.4 | 1.4-1.5 | 1.5-1.6 | 1.6-1.7 | 1.7-1.8 | 1.8-1.9 | 1.9-2.0 | 2.0-2.1 | 2.1-2.2 | 2.2-2.3 |
|---|---|---|---|---|---|---|---|---|---|---|
| **20-24** | | 101.9 [89.0, 133.1] | 110.8 [98.4, 138.0] | 121.8 [111.2, 143.3] | 131.6 [119.4, 143.8] | 148.0 [123.9, 161.5] | 149.6 [142.7, 170.8] | | | |
| **25-29** | | | 103.6 [94.9, 120.6] | 121.6 [110.3, 137.0] | 136.1 [120.4, 147.7] | 156.9 [132.4, 171.2] | 161.0 [140.2, 172.9] | 160.2 [152.5, 188.8] | | |
| **30-34** | | | 123.8 [99.0, 140.1] | 130.8 [115.1, 143.8] | 137.0 [118.8, 155.0] | 152.2 [141.5, 163.9] | 159.4 [144.3, 188.2] | 164.4 [144.4, 186.2] | 198.9 [178.8, 211.7] | |
| **35-39** | | | 109.6 [95.1, 124.8] | 132.8 [115.2, 157.6] | 139.9 [119.8, 163.1] | 150.1 [134.6, 175.5] | 156.7 [137.1, 181.5] | 174.2 [140.2, 192.8] | 202.9 [179.4, 240.8] | |
| **40-44** | | 117.0 [102.3, 129.7] | 114.1 [95.1, 134.3] | 123.2 [106.7, 144.1] | 136.2 [121.6, 158.8] | 157.5 [138.2, 181.2] | 161.3 [142.4, 186.2] | 187.1 [163.9, 209.6] | 176.2 [160.5, 191.1] | |
| **45-49** | | 76.2 [60.2, 108.7] | 118.7 [100.5, 141.8] | 124.1 [109.4, 145.0] | 141.8 [126.9, 163.6] | 149.7 [130.9, 173.6] | 165.9 [140.1, 187.3] | 191.0 [166.2, 215.5] | 200.6 [166.9, 240.8] | 195.8 [169.7, 225.7] |
| **50-54** | | 110.7 [92.4, 118.7] | 119.3 [102.7, 135.8] | 132.4 [115.3, 149.8] | 145.5 [122.4, 165.6] | 154.6 [136.8, 176.1] | 165.3 [143.7, 193.5] | 189.0 [165.6, 220.2] | 188.6 [167.1, 224.4] | 201.2 [167.1, 233.8] |
| **55-59** | | 104.3 [90.6, 120.7] | 119.3 [106.0, 138.2] | 128.8 [109.1, 145.8] | 143.7 [124.1, 163.1] | 152.3 [132.5, 175.5] | 167.2 [146.2, 191.0] | 177.4 [157.8, 196.9] | 185.1 [158.9, 221.9] | 210.6 [172.1, 238.4] |
| **60-64** | 95.1 [81.6, 101.6] | 101.9 [87.3, 113.6] | 117.5 [99.1, 134.0] | 131.9 [114.6, 148.1] | 143.3 [120.1, 162.4] | 151.9 [131.3, 171.2] | 162.8 [141.2, 188.1] | 171.4 [152.1, 195.6] | 191.1 [165.7, 212.1] | 212.1 [197.1, 231.5] |
| **65-69** | 93.8 [76.2, 110.1] | 107.2 [91.2, 126.1] | 112.8 [95.6, 132.5] | 127.3 [109.4, 146.3] | 139.1 [120.4, 157.9] | 151.5 [130.0, 170.7] | 162.1 [138.7, 186.2] | 174.7 [150.6, 195.2] | 194.8 [166.0, 214.2] | 193.6 [170.6, 223.6] |
| **70-74** | 90.7 [75.1, 102.3] | 101.7 [81.5, 117.1] | 116.4 [99.7, 134.7] | 126.8 [108.6, 145.2] | 137.3 [118.0, 156.7] | 147.4 [126.6, 168.4] | 157.6 [136.2, 181.8] | 165.7 [144.3, 189.2] | 187.8 [147.2, 242.3] | |
| **75-79** | 94.7 [80.8, 110.9] | 100.4 [82.9, 115.8] | 111.1 [94.6, 129.1] | 122.9 [104.5, 143.3] | 131.7 [114.1, 151.4] | 143.0 [122.2, 166.2] | 149.5 [132.5, 173.9] | 162.0 [142.5, 181.9] | 181.2 [165.5, 204.3] | |
| **80-84** | 86.4 [70.4, 110.1] | 95.1 [78.3, 114.1] | 108.6 [93.2, 124.5] | 118.6 [101.1, 138.1] | 129.3 [110.0, 147.9] | 135.2 [116.5, 153.0] | 142.3 [120.7, 164.3] | 157.1 [126.6, 186.1] | | |
| **85-89** | 84.7 [65.6, 95.6] | 95.1 [79.0, 108.6] | 103.7 [84.2, 120.1] | 113.2 [97.3, 131.5] | 125.6 [110.2, 143.3] | 127.7 [108.6, 154.4] | 146.3 [118.5, 168.7] | | | |

- **Female**

| | 1.1-1.2 | 1.2-1.3 | 1.3-1.4 | 1.4-1.5 | 1.5-1.6 | 1.6-1.7 | 1.7-1.8 | 1.8-1.9 | 1.9-2.0 | 2.0-2.1 |
|---|---|---|---|---|---|---|---|---|---|---|
| **20-24** | | 81.9 [60.3, 103.9] | 97.8 [88.3, 108.8] | 98.3 [90.3, 117.3] | 109.4 [97.8, 125.3] | 132.6 [118.7, 138.1] | 145.3 [127.4, 165.8] | | | |
| **25-29** | | 83.3 [55.7, 97.0] | 95.6 [83.2, 110.8] | 107.0 [93.6, 121.0] | 114.2 [101.0, 134.3] | 131.7 [113.6, 145.4] | 139.1 [129.7, 168.7] | 144.8 [134.0, 164.1] | | |
| **30-34** | | 71.8 [64.8, 83.2] | 100.0 [80.2, 111.6] | 108.9 [98.1, 121.4] | 116.3 [100.4, 132.3] | 129.9 [114.1, 153.3] | 140.0 [126.4, 148.8] | 164.9 [134.7, 180.3] | 181.3 [163.0, 196.5] | |
| **35-39** | | 95.6 [81.1, 118.4] | 96.1 [85.8, 113.1] | 107.6 [93.8, 121.7] | 117.4 [105.4, 133.9] | 125.8 [114.2, 147.3] | 141.8 [123.4, 162.0] | 154.8 [136.9, 167.9] | 157.7 [149.8, 183.8] | |
| **40-44** | | 86.9 [72.5, 94.3] | 98.2 [83.8, 111.1] | 104.3 [92.3, 117.5] | 121.7 [107.5, 134.1] | 129.6 [113.5, 145.1] | 142.0 [117.5, 159.2] | 159.8 [142.8, 184.3] | 167.8 [155.0, 185.6] | 210.5 [193.6, 225.5] |
| **45-49** | | 80.3 [70.9, 91.8] | 96.4 [83.9, 112.0] | 106.5 [92.9, 118.0] | 116.2 [103.0, 131.2] | 127.7 [111.8, 144.7] | 136.3 [122.5, 153.4] | 151.6 [133.1, 173.3] | 165.1 [152.7, 183.2] | 157.4 [132.7, 200.2] |
| **50-54** | | 83.6 [67.5, 96.1] | 95.7 [82.9, 109.3] | 104.7 [91.4, 120.3] | 117.3 [102.3, 131.6] | 126.4 [111.3, 144.0] | 141.6 [119.9, 160.8] | 154.6 [134.9, 180.2] | 168.4 [141.6, 196.9] | 165.3 [143.8, 197.7] |
| **55-59** | | 86.6 [69.1, 96.1] | 95.6 [80.6, 111.8] | 104.4 [91.3, 119.2] | 116.1 [100.8, 130.0] | 124.1 [109.4, 139.2] | 135.9 [116.3, 156.8] | 146.0 [130.2, 165.2] | 164.2 [141.5, 181.1] | |
| **60-64** | 70.0 [61.0, 83.5] | 86.7 [68.0, 96.8] | 96.1 [82.4, 111.1] | 104.9 [90.8, 119.9] | 114.1 [98.8, 130.3] | 123.5 [109.4, 144.0] | 133.2 [118.9, 151.0] | 136.2 [117.0, 155.6] | 159.1 [140.4, 184.2] | 171.4 [133.0, 178.8] |
| **65-69** | 73.5 [58.5, 95.3] | 86.7 [71.2, 99.6] | 95.5 [82.1, 110.9] | 104.7 [89.5, 116.7] | 113.5 [99.6, 128.9] | 124.4 [107.9, 142.7] | 132.8 [116.6, 150.8] | 148.4 [125.4, 172.8] | 153.0 [140.3, 163.5] | |
| **70-74** | 70.8 [57.5, 92.9] | 86.2 [68.6, 99.2] | 95.5 [81.0, 109.2] | 103.8 [90.1, 120.0] | 114.2 [97.4, 130.4] | 126.0 [104.9, 139.9] | 129.5 [111.3, 153.4] | 142.2 [118.3, 166.5] | 145.5 [127.6, 187.9] | |
| **75-79** | 68.5 [51.3, 78.0] | 81.8 [69.5, 95.7] | 94.4 [81.8, 109.1] | 101.8 [87.2, 117.6] | 111.6 [96.2, 126.7] | 121.2 [104.1, 136.1] | 127.1 [105.7, 137.6] | 133.5 [117.9, 151.8] | | |
| **80-84** | 78.4 [66.8, 89.6] | 81.6 [66.1, 94.1] | 91.1 [76.0, 106.8] | 101.5 [86.7, 117.2] | 108.2 [93.9, 127.1] | 111.8 [95.3, 126.2] | 126.3 [105.1, 147.1] | | | |
| **85-89** | 73.3 [59.4, 86.9] | 78.5 [65.6, 93.3] | 87.9 [74.1, 102.1] | 94.9 [81.2, 109.8] | 101.8 [88.2, 119.3] | 110.0 [87.4, 131.3] | 117.6 [82.7, 126.1] | | | |

### Kidney (Lt.)

#### - Male

| | 1.3-1.4 | 1.4-1.5 | 1.5-1.6 | 1.6-1.7 | 1.7-1.8 | 1.8-1.9 | 1.9-2.0 | 2.0-2.1 | 2.1-2.2 | 2.2-2.3 |
|---|---|---|---|---|---|---|---|---|---|---|
| **20-24** | | 112.9 [98.4, 126.9] | 115.0 [105.1, 140.4] | 131.3 [122.4, 157.3] | 139.1 [130.9, 151.5] | 159.6 [137.4, 171.9] | 154.9 [146.2, 185.3] | | | |
| **25-29** | | | 106.8 [68.6, 132.6] | 126.9 [103.7, 139.5] | 145.8 [129.2, 160.7] | 155.6 [141.4, 191.8] | 168.7 [147.1, 183.4] | 166.4 [150.0, 198.1] | | |
| **30-34** | | | 121.9 [108.4, 140.7] | 133.7 [117.5, 153.3] | 141.4 [125.4, 163.2] | 160.8 [145.5, 171.9] | 166.9 [145.8, 181.1] | 161.1 [154.4, 168.4] | 209.8 [174.7, 217.2] | |
| **35-39** | | | 125.7 [87.3, 136.1] | 135.4 [118.4, 157.1] | 149.1 [131.4, 168.0] | 159.2 [140.0, 178.7] | 161.4 [141.7, 182.8] | 177.2 [154.6, 199.5] | 194.1 [162.7, 222.8] | |
| **40-44** | | 125.8 [81.2, 139.4] | 126.8 [98.9, 140.7] | 136.3 [110.9, 149.9] | 140.4 [125.0, 155.2] | 160.4 [138.9, 181.3] | 172.0 [152.2, 194.7] | 192.3 [146.4, 208.3] | 188.4 [158.0, 209.6] | |
| **45-49** | | 94.9 [69.9, 117.1] | 122.0 [102.3, 146.7] | 131.5 [107.7, 151.6] | 150.1 [129.9, 174.2] | 157.4 [137.2, 178.9] | 173.2 [153.5, 198.9] | 185.8 [157.4, 207.0] | 196.5 [165.6, 226.9] | 203.5 [188.5, 248.0] |
| **50-54** | | 112.4 [90.3, 130.1] | 127.7 [111.8, 143.4] | 137.2 [117.7, 155.0] | 149.4 [125.5, 171.0] | 163.0 [137.4, 186.3] | 169.9 [148.7, 194.7] | 189.3 [165.8, 211.5] | 198.0 [174.7, 227.6] | 213.4 [185.1, 246.6] |
| **55-59** | | 115.8 [90.1, 133.7] | 130.3 [104.4, 145.6] | 137.4 [118.7, 156.9] | 148.6 [129.6, 171.1] | 159.1 [136.9, 183.6] | 171.5 [148.4, 195.7] | 183.3 [160.8, 205.5] | 192.8 [162.5, 222.3] | 202.6 [167.6, 235.3] |
| **60-64** | 86.4 [80.5, 112.0] | 109.5 [98.3, 127.7] | 123.3 [106.1, 139.0] | 136.7 [120.2, 153.6] | 146.0 [124.5, 167.8] | 155.8 [135.5, 177.5] | 166.1 [145.0, 190.3] | 174.1 [150.2, 196.5] | 197.1 [169.0, 219.0] | 215.0 [188.5, 225.3] |
| **65-69** | 93.5 [72.5, 111.4] | 113.1 [96.1, 132.0] | 118.6 [100.7, 137.2] | 133.0 [114.1, 152.3] | 141.8 [120.4, 162.9] | 153.6 [132.9, 174.6] | 165.4 [142.1, 189.5] | 172.5 [147.7, 195.3] | 188.7 [165.4, 213.7] | 189.5 [180.1, 215.0] |
| **70-74** | 95.0 [78.8, 113.2] | 109.5 [88.8, 128.6] | 121.8 [103.4, 139.4] | 130.9 [110.5, 150.4] | 141.9 [121.1, 162.0] | 152.1 [130.4, 175.8] | 160.0 [136.4, 183.8] | 169.1 [146.8, 194.2] | 174.7 [135.8, 220.3] | |
| **75-79** | 95.8 [81.5, 117.7] | 105.8 [86.6, 124.9] | 116.2 [96.0, 134.6] | 127.8 [108.7, 147.8] | 135.8 [116.9, 158.0] | 148.7 [126.4, 170.6] | 156.2 [132.8, 177.3] | 166.4 [136.6, 191.2] | 171.0 [150.9, 193.1] | |
| **80-84** | 87.0 [70.5, 106.8] | 100.7 [82.7, 122.8] | 113.3 [95.0, 131.4] | 121.5 [100.4, 140.9] | 133.5 [114.3, 154.9] | 139.1 [116.6, 159.6] | 145.3 [123.4, 174.3] | 158.2 [147.5, 177.8] | | |
| **85-89** | 82.1 [57.0, 103.4] | 94.0 [79.3, 110.6] | 105.5 [87.8, 127.6] | 117.8 [99.6, 140.5] | 129.0 [107.4, 153.5] | 129.2 [109.5, 151.2] | 149.0 [126.3, 172.3] | | | |

#### - Female

| | 1.1-1.2 | 1.2-1.3 | 1.3-1.4 | 1.4-1.5 | 1.5-1.6 | 1.6-1.7 | 1.7-1.8 | 1.8-1.9 | 1.9-2.0 | 2.0-2.1 |
|---|---|---|---|---|---|---|---|---|---|---|
| **20-24** | | 69.3 [46.3, 103.3] | 107.4 [94.4, 117.5] | 106.0 [86.0, 132.0] | 118.2 [104.5, 136.4] | 133.4 [121.4, 145.5] | 145.5 [127.0, 163.8] | | | |
| **25-29** | | 84.1 [65.2, 104.7] | 106.9 [89.8, 114.8] | 112.0 [98.6, 126.5] | 124.1 [115.6, 141.8] | 137.7 [126.8, 151.7] | 157.0 [137.4, 176.3] | 156.2 [139.8, 169.5] | | |
| **30-34** | | 76.8 [49.8, 97.3] | 107.8 [84.2, 124.0] | 113.9 [104.6, 130.4] | 127.9 [110.6, 145.6] | 138.4 [124.8, 167.9] | 149.4 [131.0, 165.1] | 150.6 [142.3, 198.2] | 191.5 [185.7, 200.7] | |
| **35-39** | | 98.2 [80.9, 131.2] | 102.2 [87.3, 117.4] | 111.2 [96.6, 125.4] | 123.8 [107.3, 140.7] | 135.1 [121.7, 156.2] | 154.2 [128.6, 176.5] | 161.9 [140.9, 174.2] | 169.6 [154.1, 192.4] | |
| **40-44** | | 86.5 [70.6, 101.2] | 99.5 [87.0, 114.2] | 109.5 [95.3, 126.7] | 125.9 [112.5, 142.2] | 134.3 [118.6, 155.1] | 149.8 [125.0, 170.0] | 164.8 [142.8, 183.2] | 172.6 [162.6, 192.6] | 211.5 [200.9, 256.2] |
| **45-49** | | 84.3 [23.9, 97.3] | 103.4 [87.8, 117.9] | 111.1 [99.1, 127.4] | 124.3 [110.3, 141.4] | 135.1 [119.0, 151.5] | 146.0 [128.9, 160.4] | 164.5 [145.3, 177.8] | 167.4 [151.2, 178.2] | 179.5 [143.0, 217.9] |
| **50-54** | | 83.7 [62.5, 107.1] | 101.8 [86.3, 114.8] | 112.9 [96.7, 127.8] | 123.9 [109.0, 139.6] | 133.3 [117.6, 151.4] | 149.6 [127.2, 169.5] | 157.5 [136.7, 178.2] | 169.7 [150.4, 198.3] | 183.1 [143.5, 212.3] |
| **55-59** | | 84.3 [55.0, 106.7] | 102.6 [87.0, 116.0] | 109.1 [96.3, 125.3] | 120.7 [104.0, 134.9] | 130.7 [112.5, 146.2] | 145.1 [121.5, 162.1] | 153.0 [131.0, 165.2] | 167.5 [144.6, 189.2] | |
| **60-64** | 71.5 [26.5, 83.3] | 92.4 [71.8, 105.7] | 99.1 [82.7, 116.4] | 113.0 [97.5, 128.5] | 121.2 [104.3, 138.1] | 129.3 [114.4, 147.7] | 139.6 [117.3, 156.5] | 143.3 [121.9, 165.4] | 163.1 [149.4, 193.4] | 177.3 [155.6, 252.4] |
| **65-69** | 84.5 [73.2, 100.6] | 93.8 [72.0, 106.8] | 100.7 [86.1, 116.6] | 111.3 [96.2, 126.2] | 120.6 [105.7, 136.3] | 129.6 [112.0, 148.1] | 135.3 [116.5, 150.8] | 143.3 [124.1, 173.9] | 146.3 [139.8, 163.9] | |
| **70-74** | 74.7 [47.9, 86.9] | 91.0 [73.2, 109.3] | 100.4 [83.9, 115.4] | 108.5 [92.7, 127.3] | 118.9 [102.2, 136.3] | 127.8 [107.9, 148.3] | 135.7 [113.5, 160.1] | 153.8 [121.3, 176.5] | 129.9 [122.0, 139.8] | |
| **75-79** | 76.0 [58.7, 87.4] | 90.0 [74.6, 106.1] | 99.1 [83.7, 113.8] | 108.4 [91.3, 125.9] | 116.3 [100.6, 132.4] | 124.1 [106.1, 143.9] | 125.1 [106.2, 148.7] | 123.0 [86.7, 141.9] | | |
| **80-84** | 79.7 [67.4, 94.3] | 85.4 [66.3, 100.6] | 96.6 [81.4, 113.3] | 105.7 [90.0, 123.8] | 113.1 [96.8, 132.3] | 117.1 [103.8, 132.3] | 130.1 [111.1, 145.6] | | | |
| **85-89** | 74.6 [59.0, 96.1] | 81.6 [68.2, 96.9] | 93.8 [79.4, 108.8] | 100.1 [83.2, 113.9] | 108.8 [88.9, 126.9] | 114.1 [89.9, 135.1] | 125.0 [93.8, 134.3] | | | |

### Kidney (Total)

#### - Male

| | 1.3-1.4 | 1.4-1.5 | 1.5-1.6 | 1.6-1.7 | 1.7-1.8 | 1.8-1.9 | 1.9-2.0 | 2.0-2.1 | 2.1-2.2 | 2.2-2.3 |
|---|---|---|---|---|---|---|---|---|---|---|
| **20-24** | | 213.6<br>[190.8, 262.4] | 234.6<br>[200.5, 272.3] | 253.1<br>[229.8, 290.9] | 268.2<br>[257.3, 293.5] | 309.2<br>[268.2, 334.6] | 304.5<br>[288.9, 356.1] | | | |
| **25-29** | | | 207.2<br>[166.6, 252.9] | 252.8<br>[209.5, 277.0] | 279.4<br>[258.4, 303.8] | 316.8<br>[276.5, 360.8] | 317.4<br>[289.4, 345.1] | 321.7<br>[296.4, 385.3] | | |
| **30-34** | | | 247.2<br>[210.7, 281.9] | 265.7<br>[228.4, 289.5] | 278.7<br>[246.6, 307.3] | 311.7<br>[286.6, 332.9] | 327.5<br>[292.1, 368.0] | 324.6<br>[304.1, 363.6] | 407.5<br>[344.6, 424.5] | |
| **35-39** | | | 234.7<br>[192.1, 258.9] | 271.6<br>[235.3, 305.5] | 286.7<br>[254.8, 329.5] | 303.7<br>[280.3, 358.7] | 315.6<br>[281.1, 354.2] | 337.0<br>[294.8, 385.9] | 385.6<br>[344.5, 438.8] | |
| **40-44** | | 242.8<br>[183.4, 270.1] | 246.2<br>[197.9, 277.1] | 262.2<br>[220.1, 295.8] | 276.8<br>[252.0, 310.0] | 315.1<br>[276.3, 355.7] | 333.0<br>[300.4, 383.7] | 369.5<br>[321.3, 410.5] | 367.3<br>[325.0, 394.5] | |
| **45-49** | | 182.8<br>[125.8, 220.8] | 234.5<br>[198.0, 281.0] | 255.7<br>[207.7, 294.6] | 290.4<br>[261.0, 328.7] | 306.9<br>[268.3, 346.1] | 331.9<br>[286.2, 379.7] | 369.3<br>[328.3, 401.6] | 400.7<br>[351.4, 476.4] | 396.3<br>[360.6, 469.0] |
| **50-54** | | 222.7<br>[175.6, 245.1] | 243.5<br>[214.5, 267.6] | 265.1<br>[234.7, 302.6] | 291.8<br>[248.5, 331.2] | 312.7<br>[272.0, 351.2] | 332.3<br>[289.1, 375.2] | 374.1<br>[327.4, 419.4] | 387.4<br>[341.4, 436.4] | 400.3<br>[345.0, 446.7] |
| **55-59** | | 214.0<br>[180.9, 249.7] | 248.3<br>[212.9, 285.6] | 264.0<br>[228.3, 298.6] | 289.8<br>[253.5, 332.0] | 308.7<br>[268.0, 350.7] | 337.9<br>[292.8, 375.2] | 357.5<br>[320.3, 395.4] | 367.9<br>[325.7, 428.0] | 407.6<br>[351.2, 437.4] |
| **60-64** | 181.5<br>[145.9, 212.6] | 211.7<br>[191.0, 236.9] | 237.4<br>[210.6, 271.7] | 265.3<br>[232.7, 299.5] | 285.3<br>[245.1, 321.3] | 305.6<br>[265.0, 345.1] | 328.7<br>[287.6, 370.5] | 344.7<br>[300.2, 383.5] | 371.1<br>[337.7, 414.4] | 431.6<br>[391.4, 448.1] |
| **65-69** | 183.9<br>[156.1, 217.5] | 215.1<br>[188.1, 249.0] | 230.8<br>[195.0, 263.8] | 255.6<br>[221.2, 290.5] | 278.3<br>[239.6, 317.0] | 300.6<br>[261.9, 339.2] | 321.4<br>[277.9, 368.9] | 345.7<br>[296.8, 391.0] | 365.5<br>[331.2, 444.9] | 376.5<br>[350.3, 430.9] |
| **70-74** | 182.8<br>[152.0, 206.2] | 208.7<br>[172.9, 241.1] | 237.3<br>[200.9, 271.2] | 254.7<br>[220.7, 291.5] | 273.9<br>[236.1, 311.9] | 295.1<br>[255.2, 337.4] | 315.2<br>[273.4, 358.5] | 332.5<br>[291.5, 379.7] | 343.8<br>[295.1, 444.9] | |
| **75-79** | 187.7<br>[169.8, 224.6] | 203.5<br>[170.0, 234.4] | 225.6<br>[190.3, 258.6] | 246.8<br>[212.4, 284.0] | 264.9<br>[230.3, 302.9] | 287.0<br>[248.8, 330.9] | 303.5<br>[261.2, 342.1] | 328.8<br>[276.0, 370.5] | 346.1<br>[318.8, 396.0] | |
| **80-84** | 170.0<br>[146.0, 206.6] | 194.9<br>[165.2, 226.5] | 220.6<br>[186.2, 252.2] | 235.9<br>[203.0, 272.7] | 258.9<br>[222.1, 294.7] | 268.6<br>[229.2, 309.4] | 276.6<br>[242.6, 334.1] | 309.4<br>[269.5, 370.8] | | |
| **85-89** | 159.8<br>[124.5, 196.7] | 185.9<br>[159.5, 216.8] | 208.7<br>[174.7, 242.8] | 229.7<br>[194.1, 267.4] | 253.0<br>[215.9, 290.8] | 253.7<br>[215.6, 287.6] | 294.4<br>[241.8, 331.2] | | | |

#### - Female

| | 1.1-1.2 | 1.2-1.3 | 1.3-1.4 | 1.4-1.5 | 1.5-1.6 | 1.6-1.7 | 1.7-1.8 | 1.8-1.9 | 1.9-2.0 | 2.0-2.1 |
|---|---|---|---|---|---|---|---|---|---|---|
| **20-24** | | 167.9<br>[131.0, 208.1] | 204.9<br>[179.5, 220.1] | 202.9<br>[181.2, 240.7] | 222.3<br>[208.4, 259.3] | 264.9<br>[245.1, 285.8] | 285.6<br>[253.5, 320.6] | | | |
| **25-29** | | 166.3<br>[122.3, 205.1] | 200.0<br>[180.4, 223.8] | 218.8<br>[193.4, 243.7] | 238.2<br>[218.5, 272.4] | 270.5<br>[242.8, 298.5] | 315.3<br>[268.2, 327.7] | 303.9<br>[273.7, 320.3] | | |
| **30-34** | | 150.0<br>[93.9, 171.3] | 207.6<br>[178.5, 235.6] | 224.0<br>[202.6, 245.4] | 243.7<br>[209.9, 274.7] | 269.1<br>[243.6, 315.2] | 286.9<br>[258.1, 312.1] | 309.2<br>[275.6, 379.3] | 372.9<br>[348.7, 397.2] | |
| **35-39** | | 192.8<br>[159.3, 249.2] | 196.2<br>[171.2, 226.9] | 219.4<br>[190.3, 242.8] | 240.2<br>[213.7, 270.2] | 259.9<br>[233.8, 300.7] | 300.5<br>[248.9, 337.6] | 318.3<br>[280.9, 340.3] | 322.7<br>[307.3, 375.2] | |
| **40-44** | | 163.1<br>[154.3, 185.1] | 197.7<br>[175.1, 224.9] | 214.7<br>[185.8, 238.1] | 244.2<br>[221.2, 276.3] | 260.9<br>[232.2, 298.8] | 283.0<br>[240.5, 327.7] | 329.1<br>[285.7, 359.9] | 341.0<br>[317.7, 384.3] | 422.2<br>[391.7, 494.2] |
| **45-49** | | 157.4<br>[119.8, 185.3] | 200.7<br>[169.8, 224.3] | 213.6<br>[194.2, 244.3] | 240.2<br>[214.1, 267.2] | 260.0<br>[233.8, 294.4] | 273.9<br>[252.0, 313.0] | 311.4<br>[281.1, 341.3] | 331.8<br>[304.6, 354.8] | 337.0<br>[278.7, 425.4] |
| **50-54** | | 162.5<br>[132.8, 205.0] | 195.7<br>[169.9, 221.5] | 216.4<br>[188.8, 242.1] | 238.4<br>[212.4, 267.6] | 257.8<br>[229.5, 287.4] | 290.2<br>[251.7, 323.5] | 310.0<br>[268.9, 352.4] | 336.2<br>[295.2, 394.5] | 356.4<br>[297.4, 409.3] |
| **55-59** | | 168.5<br>[132.7, 203.7] | 198.4<br>[165.9, 224.4] | 211.9<br>[188.6, 241.0] | 236.2<br>[206.3, 261.2] | 253.0<br>[221.7, 283.0] | 277.5<br>[239.3, 314.2] | 300.5<br>[267.6, 328.6] | 336.1<br>[299.3, 355.1] | |
| **60-64** | 144.6<br>[83.2, 161.1] | 170.9<br>[134.5, 200.5] | 195.2<br>[168.5, 224.7] | 215.0<br>[186.7, 244.2] | 233.3<br>[203.5, 262.2] | 250.5<br>[222.3, 285.2] | 272.2<br>[238.4, 307.2] | 280.9<br>[240.2, 317.8] | 308.8<br>[282.7, 371.8] | 342.9<br>[259.9, 366.8] |
| **65-69** | 168.9<br>[128.2, 181.9] | 177.5<br>[150.8, 201.7] | 193.8<br>[168.0, 220.6] | 214.8<br>[186.9, 240.0] | 232.5<br>[204.8, 260.8] | 253.4<br>[220.4, 287.7] | 266.1<br>[233.5, 299.1] | 295.1<br>[253.3, 338.7] | 297.1<br>[275.0, 327.2] | |
| **70-74** | 146.8<br>[97.0, 169.7] | 172.1<br>[142.1, 201.1] | 192.6<br>[164.8, 221.3] | 210.3<br>[181.7, 242.0] | 230.6<br>[201.1, 262.0] | 251.2<br>[211.3, 283.2] | 259.2<br>[224.8, 307.0] | 291.6<br>[247.8, 337.7] | 268.2<br>[248.7, 316.9] | |
| **75-79** | 144.9<br>[118.9, 161.5] | 170.4<br>[142.2, 199.1] | 192.5<br>[165.1, 219.0] | 206.7<br>[176.8, 239.4] | 227.0<br>[196.5, 256.6] | 240.8<br>[206.3, 277.0] | 250.1<br>[211.3, 285.1] | 260.5<br>[190.2, 281.8] | | |
| **80-84** | 148.8<br>[133.8, 170.0] | 161.6<br>[132.4, 188.3] | 185.0<br>[157.8, 214.0] | 204.9<br>[176.2, 233.3] | 220.3<br>[188.2, 252.6] | 228.8<br>[196.7, 260.0] | 241.9<br>[216.5, 291.7] | | | |
| **85-89** | 144.1<br>[123.5, 174.9] | 160.8<br>[135.8, 185.7] | 182.1<br>[153.7, 207.0] | 192.6<br>[164.1, 218.2] | 206.7<br>[175.4, 243.4] | 221.3<br>[178.9, 254.2] | 242.8<br>[176.5, 272.3] | | | |

### Adrenal gland (Rt.)

- **Male**

| | **1.3-1.4** | **1.4-1.5** | **1.5-1.6** | **1.6-1.7** | **1.7-1.8** | **1.8-1.9** | **1.9-2.0** | **2.0-2.1** | **2.1-2.2** | **2.2-2.3** |
|---|---|---|---|---|---|---|---|---|---|---|
| **20-24** | | 1.1 [0.8, 1.7] | 1.2 [0.6, 1.8] | 1.4 [0.9, 1.8] | 1.6 [1.1, 1.8] | 1.8 [1.5, 2.6] | 2.3 [1.9, 2.7] | | | |
| **25-29** | | | 1.2 [0.9, 1.9] | 1.5 [1.0, 2.1] | 1.9 [1.5, 2.5] | 2.5 [1.9, 3.2] | 2.6 [2.0, 2.8] | 2.9 [1.9, 3.9] | | |
| **30-34** | | | 1.7 [1.3, 1.9] | 1.8 [1.3, 2.4] | 2.0 [1.5, 2.7] | 2.1 [1.5, 2.5] | 2.6 [2.2, 3.3] | 2.7 [2.1, 3.2] | 2.8 [2.6, 4.0] | |
| **35-39** | | | 1.7 [1.3, 2.2] | 2.0 [1.4, 2.7] | 2.1 [1.6, 2.6] | 2.3 [1.7, 3.0] | 2.8 [2.3, 3.5] | 3.2 [2.4, 3.8] | 4.2 [3.2, 5.1] | |
| **40-44** | | 1.8 [1.5, 2.3] | 1.5 [1.0, 1.9] | 2.1 [1.5, 2.7] | 2.3 [1.7, 3.0] | 2.8 [2.2, 3.3] | 3.3 [2.5, 3.8] | 3.4 [2.9, 4.0] | 3.8 [2.9, 4.3] | |
| **45-49** | | 1.6 [0.7, 2.0] | 2.1 [1.4, 2.6] | 2.1 [1.5, 2.7] | 2.6 [2.1, 3.4] | 2.9 [2.2, 3.7] | 3.2 [2.5, 3.8] | 3.8 [3.2, 4.7] | 4.1 [3.6, 4.8] | 4.5 [4.0, 5.2] |
| **50-54** | | 1.7 [1.3, 2.4] | 2.1 [1.3, 2.8] | 2.4 [1.8, 3.1] | 2.8 [2.1, 3.4] | 3.0 [2.3, 3.7] | 3.3 [2.7, 4.1] | 3.9 [3.0, 4.6] | 4.1 [3.3, 4.9] | 4.9 [4.0, 5.6] |
| **55-59** | | 1.9 [1.3, 2.5] | 2.2 [1.4, 2.8] | 2.6 [1.8, 3.2] | 2.9 [2.3, 3.6] | 3.3 [2.6, 3.9] | 3.6 [2.7, 4.4] | 3.9 [3.0, 4.6] | 4.2 [3.3, 4.7] | 4.7 [4.1, 5.3] |
| **60-64** | 1.4 [1.3, 1.9] | 1.9 [1.3, 2.6] | 2.3 [1.8, 3.0] | 2.7 [2.0, 3.3] | 3.0 [2.4, 3.7] | 3.4 [2.7, 4.0] | 3.6 [2.9, 4.3] | 3.7 [3.0, 4.4] | 4.3 [3.3, 5.5] | 4.9 [3.7, 5.5] |
| **65-69** | 1.5 [0.8, 2.8] | 1.9 [1.3, 2.5] | 2.4 [1.6, 3.0] | 2.8 [2.2, 3.4] | 3.1 [2.5, 3.7] | 3.4 [2.8, 4.0] | 3.7 [3.0, 4.3] | 3.8 [3.2, 4.5] | 4.5 [3.9, 5.2] | 4.2 [3.6, 4.7] |
| **70-74** | 1.6 [1.0, 2.2] | 1.9 [1.2, 2.6] | 2.5 [1.8, 3.1] | 2.8 [2.2, 3.5] | 3.1 [2.6, 3.8] | 3.5 [2.9, 4.1] | 3.7 [3.1, 4.4] | 4.0 [3.3, 4.8] | 4.5 [3.6, 5.2] | |
| **75-79** | 1.7 [1.0, 2.5] | 2.1 [1.2, 2.9] | 2.5 [1.9, 3.2] | 2.9 [2.3, 3.6] | 3.2 [2.6, 3.8] | 3.4 [2.9, 4.1] | 3.6 [3.0, 4.4] | 3.9 [3.3, 4.5] | 4.0 [3.6, 5.0] | |
| **80-84** | 1.9 [0.9, 2.8] | 2.1 [1.3, 2.8] | 2.6 [2.0, 3.2] | 3.0 [2.4, 3.6] | 3.1 [2.5, 3.8] | 3.2 [2.7, 3.8] | 3.5 [2.9, 4.3] | 3.9 [3.0, 4.9] | | |
| **85-89** | 2.1 [1.0, 2.6] | 2.4 [1.5, 3.0] | 2.6 [1.9, 3.2] | 2.9 [2.3, 3.4] | 3.2 [2.5, 3.9] | 3.3 [2.8, 4.0] | 3.6 [3.0, 4.3] | | | |

- **Female**

| | **1.1-1.2** | **1.2-1.3** | **1.3-1.4** | **1.4-1.5** | **1.5-1.6** | **1.6-1.7** | **1.7-1.8** | **1.8-1.9** | **1.9-2.0** | **2.0-2.1** |
|---|---|---|---|---|---|---|---|---|---|---|
| **20-24** | | 0.5 [0.3, 0.9] | 0.9 [0.6, 1.4] | 0.8 [0.5, 1.2] | 1.0 [0.7, 1.3] | 1.2 [0.7, 1.7] | 1.3 [0.9, 1.7] | | | |
| **25-29** | | 1.0 [0.5, 1.2] | 1.0 [0.6, 1.4] | 0.9 [0.5, 1.2] | 1.3 [0.9, 1.6] | 1.4 [0.9, 1.6] | 1.8 [1.2, 2.3] | 1.7 [1.5, 2.2] | | |
| **30-34** | | 1.1 [0.8, 1.7] | 1.0 [0.6, 1.4] | 0.9 [0.6, 1.4] | 1.1 [0.8, 1.4] | 1.4 [1.0, 1.9] | 1.8 [1.3, 2.3] | 1.7 [1.3, 2.4] | 2.5 [2.1, 2.6] | |
| **35-39** | | 1.3 [0.7, 1.7] | 1.0 [0.7, 1.3] | 1.1 [0.7, 1.5] | 1.3 [0.9, 1.8] | 1.6 [1.3, 2.1] | 2.0 [1.4, 2.4] | 2.4 [1.7, 2.7] | 2.5 [1.7, 2.9] | |
| **40-44** | | 0.5 [0.4, 1.1] | 1.0 [0.6, 1.5] | 1.2 [0.8, 1.6] | 1.4 [1.0, 1.8] | 1.6 [1.2, 2.1] | 2.2 [1.5, 2.7] | 2.5 [1.8, 3.2] | 3.0 [2.6, 3.4] | 3.7 [2.9, 4.4] |
| **45-49** | | 0.9 [0.4, 1.2] | 1.0 [0.6, 1.4] | 1.3 [0.9, 1.7] | 1.5 [1.1, 2.0] | 1.8 [1.3, 2.3] | 2.2 [1.7, 2.9] | 2.8 [2.4, 3.2] | 3.3 [2.8, 3.9] | 2.9 [2.6, 3.4] |
| **50-54** | | 0.9 [0.5, 1.4] | 1.2 [0.7, 1.6] | 1.4 [1.0, 1.8] | 1.6 [1.2, 2.1] | 2.1 [1.5, 2.6] | 2.5 [1.9, 3.2] | 2.8 [2.2, 3.6] | 3.1 [2.6, 3.8] | 3.5 [2.6, 3.8] |
| **55-59** | | 1.0 [0.3, 1.4] | 1.1 [0.7, 1.6] | 1.4 [0.9, 1.9] | 1.8 [1.3, 2.3] | 2.2 [1.6, 2.6] | 2.7 [2.2, 3.4] | 2.9 [2.4, 3.5] | 3.2 [2.9, 3.7] | |
| **60-64** | 1.0 [0.6, 1.7] | 1.0 [0.5, 1.5] | 1.3 [0.8, 1.7] | 1.6 [1.1, 2.1] | 1.9 [1.4, 2.5] | 2.3 [1.8, 3.0] | 2.8 [2.3, 3.3] | 2.9 [2.3, 3.5] | 3.6 [2.8, 3.9] | 3.6 [3.4, 4.3] |
| **65-69** | 0.7 [0.2, 1.5] | 1.2 [0.7, 1.6] | 1.5 [0.9, 1.9] | 1.8 [1.1, 2.4] | 2.1 [1.6, 2.8] | 2.5 [2.0, 3.1] | 2.9 [2.4, 3.5] | 3.4 [2.6, 3.9] | 3.4 [3.0, 4.1] | |
| **70-74** | 0.9 [0.4, 1.5] | 1.1 [0.5, 1.7] | 1.5 [1.0, 2.2] | 1.9 [1.4, 2.5] | 2.3 [1.7, 2.9] | 2.8 [2.2, 3.4] | 2.9 [2.3, 3.5] | 3.3 [2.5, 3.6] | 3.3 [2.4, 4.1] | |
| **75-79** | 1.1 [0.3, 1.8] | 1.3 [0.7, 1.9] | 1.7 [1.1, 2.3] | 2.1 [1.5, 2.6] | 2.4 [1.9, 3.1] | 2.8 [2.2, 3.5] | 2.9 [2.5, 3.5] | 3.2 [2.5, 3.9] | | |
| **80-84** | 1.1 [0.7, 1.5] | 1.4 [0.7, 2.0] | 1.8 [1.2, 2.4] | 2.2 [1.7, 2.7] | 2.5 [1.9, 3.1] | 2.9 [2.4, 3.4] | 3.2 [2.3, 3.8] | | | |
| **85-89** | 1.0 [0.3, 1.8] | 1.4 [0.6, 2.0] | 1.9 [1.4, 2.5] | 2.2 [1.8, 2.8] | 2.5 [2.0, 3.1] | 2.8 [2.1, 3.3] | 3.5 [2.9, 4.5] | | | |

### Adrenal gland (Lt.)

#### - Male

| | 1.3-1.4 | 1.4-1.5 | 1.5-1.6 | 1.6-1.7 | 1.7-1.8 | 1.8-1.9 | 1.9-2.0 | 2.0-2.1 | 2.1-2.2 | 2.2-2.3 |
|---|---|---|---|---|---|---|---|---|---|---|
| **20-24** | | 1.7 [0.9, 2.4] | 1.5 [1.0, 2.5] | 1.9 [1.1, 2.6] | 2.1 [1.5, 2.4] | 2.4 [1.5, 3.1] | 3.3 [2.9, 3.4] | | | |
| **25-29** | | | 1.8 [1.2, 2.5] | 2.1 [1.4, 2.8] | 2.5 [2.0, 3.3] | 3.1 [2.5, 4.3] | 2.9 [2.1, 3.4] | 3.7 [2.8, 4.3] | | |
| **30-34** | | | 2.2 [1.7, 2.5] | 2.4 [1.9, 3.0] | 2.7 [2.0, 3.4] | 3.2 [2.3, 3.7] | 3.6 [2.7, 4.1] | 3.7 [3.2, 4.5] | 4.6 [4.1, 5.0] | |
| **35-39** | | | 2.0 [0.9, 2.8] | 2.5 [1.8, 3.5] | 3.1 [2.3, 3.7] | 3.2 [2.4, 4.0] | 3.6 [2.6, 4.5] | 4.0 [3.3, 4.6] | 4.5 [4.0, 5.2] | |
| **40-44** | | 2.5 [0.7, 3.1] | 2.3 [1.5, 2.9] | 2.7 [1.9, 3.5] | 3.1 [2.3, 3.8] | 3.7 [2.9, 4.4] | 4.1 [3.6, 4.9] | 4.8 [3.7, 5.6] | 4.8 [3.9, 5.6] | |
| **45-49** | | 1.7 [1.2, 2.7] | 2.6 [1.9, 3.5] | 2.8 [1.9, 3.4] | 3.4 [2.8, 4.2] | 3.6 [2.9, 4.4] | 4.0 [3.3, 4.9] | 4.3 [3.8, 5.6] | 4.9 [3.9, 6.1] | 6.1 [5.0, 7.0] |
| **50-54** | | 2.6 [1.7, 3.1] | 2.8 [2.0, 3.4] | 3.1 [2.3, 3.8] | 3.5 [2.7, 4.2] | 3.9 [3.1, 4.9] | 4.5 [3.5, 5.2] | 4.9 [3.9, 5.8] | 5.4 [4.6, 6.0] | 5.9 [5.0, 6.7] |
| **55-59** | | 2.3 [1.3, 2.8] | 2.8 [2.1, 3.8] | 3.3 [2.6, 4.2] | 3.8 [2.9, 4.5] | 4.2 [3.3, 5.0] | 4.6 [3.5, 5.5] | 4.7 [4.0, 5.7] | 5.3 [4.3, 6.2] | 5.7 [5.0, 7.1] |
| **60-64** | 2.0 [1.7, 2.6] | 2.5 [1.7, 3.6] | 3.0 [2.0, 3.8] | 3.5 [2.7, 4.2] | 3.8 [3.1, 4.6] | 4.3 [3.5, 5.2] | 4.6 [3.8, 5.6] | 4.8 [3.9, 5.9] | 5.3 [4.0, 6.5] | 6.0 [5.3, 7.5] |
| **65-69** | 1.9 [0.6, 3.0] | 2.5 [1.8, 3.3] | 3.0 [1.9, 3.8] | 3.5 [2.7, 4.2] | 3.9 [3.1, 4.7] | 4.2 [3.5, 5.2] | 4.6 [3.7, 5.5] | 4.9 [3.9, 6.1] | 5.3 [4.8, 6.9] | 4.9 [4.5, 6.3] |
| **70-74** | 2.1 [1.1, 2.6] | 2.6 [1.6, 3.4] | 3.1 [2.2, 3.8] | 3.5 [2.6, 4.3] | 3.9 [3.2, 4.8] | 4.3 [3.6, 5.3] | 4.7 [3.8, 5.5] | 4.9 [4.3, 5.6] | 5.1 [4.0, 5.8] | |
| **75-79** | 2.1 [1.5, 2.9] | 2.7 [1.7, 3.5] | 3.1 [2.2, 4.0] | 3.7 [2.9, 4.5] | 4.0 [3.2, 4.8] | 4.3 [3.5, 5.1] | 4.6 [3.7, 5.6] | 5.1 [4.1, 6.2] | 5.2 [4.5, 7.1] | |
| **80-84** | 2.2 [0.9, 3.0] | 2.8 [1.7, 3.5] | 3.2 [2.3, 4.0] | 3.6 [2.8, 4.4] | 4.0 [3.2, 4.8] | 4.2 [3.5, 5.0] | 4.3 [3.6, 5.1] | 4.6 [4.0, 5.2] | | |
| **85-89** | 2.5 [1.2, 3.2] | 2.7 [1.7, 3.6] | 3.1 [2.3, 3.9] | 3.5 [2.8, 4.2] | 3.9 [3.0, 4.7] | 4.2 [3.4, 4.8] | 4.9 [4.0, 5.3] | | | |

#### - Female

| | 1.1-1.2 | 1.2-1.3 | 1.3-1.4 | 1.4-1.5 | 1.5-1.6 | 1.6-1.7 | 1.7-1.8 | 1.8-1.9 | 1.9-2.0 | 2.0-2.1 |
|---|---|---|---|---|---|---|---|---|---|---|
| **20-24** | | 1.1 [0.3, 1.8] | 1.2 [0.8, 1.5] | 1.2 [0.7, 1.7] | 1.5 [0.8, 1.8] | 1.6 [0.9, 2.2] | 2.0 [1.6, 2.6] | | | |
| **25-29** | | 0.7 [0.5, 1.6] | 1.2 [0.8, 1.6] | 1.3 [0.7, 1.8] | 1.7 [1.2, 2.4] | 1.8 [1.2, 2.3] | 2.6 [2.1, 3.3] | 2.4 [1.9, 2.9] | | |
| **30-34** | | 1.1 [0.7, 1.5] | 1.7 [1.2, 2.2] | 1.6 [1.0, 2.0] | 1.7 [1.1, 2.2] | 2.0 [1.5, 2.5] | 2.7 [1.8, 3.3] | 2.9 [2.4, 3.4] | 3.2 [2.3, 4.2] | |
| **35-39** | | 1.9 [1.2, 2.2] | 1.2 [0.4, 1.8] | 1.6 [1.0, 2.2] | 1.8 [1.2, 2.2] | 2.1 [1.6, 2.6] | 2.7 [2.0, 3.1] | 2.9 [2.5, 3.5] | 3.4 [2.9, 4.1] | |
| **40-44** | | 0.5 [0.2, 1.3] | 1.5 [0.9, 1.9] | 1.6 [0.9, 2.1] | 2.0 [1.4, 2.6] | 2.3 [1.6, 2.9] | 2.8 [2.2, 3.5] | 3.0 [2.5, 3.7] | 3.8 [3.3, 4.5] | 4.9 [4.1, 5.2] |
| **45-49** | | 1.2 [0.2, 1.7] | 1.4 [0.9, 1.9] | 1.7 [1.1, 2.4] | 2.2 [1.5, 2.7] | 2.4 [1.8, 3.1] | 2.9 [2.3, 3.6] | 3.6 [2.9, 4.4] | 4.0 [3.4, 5.0] | 3.6 [3.2, 4.6] |
| **50-54** | | 1.3 [0.5, 2.3] | 1.6 [0.9, 2.2] | 1.8 [1.2, 2.5] | 2.2 [1.6, 2.8] | 2.6 [1.9, 3.4] | 3.2 [2.3, 3.9] | 3.6 [3.0, 4.2] | 3.8 [3.0, 4.6] | 4.6 [3.9, 5.5] |
| **55-59** | | 0.8 [0.0, 2.0] | 1.5 [0.8, 2.3] | 2.0 [1.2, 2.6] | 2.3 [1.7, 3.0] | 2.8 [2.2, 3.5] | 3.5 [2.7, 4.2] | 3.6 [2.8, 4.5] | 4.4 [3.6, 4.9] | |
| **60-64** | 0.9 [0.0, 1.6] | 1.2 [0.7, 2.0] | 1.6 [0.9, 2.3] | 2.1 [1.5, 2.8] | 2.6 [1.9, 3.3] | 3.0 [2.3, 3.8] | 3.4 [2.8, 4.0] | 3.8 [2.9, 4.6] | 4.3 [3.8, 5.1] | 4.4 [3.6, 6.1] |
| **65-69** | 1.0 [0.4, 1.5] | 1.4 [0.6, 2.1] | 1.8 [0.9, 2.5] | 2.3 [1.6, 3.0] | 2.8 [2.1, 3.5] | 3.1 [2.5, 3.8] | 3.6 [3.0, 4.3] | 3.9 [3.1, 5.0] | 4.6 [3.9, 5.4] | |
| **70-74** | 0.8 [0.1, 2.0] | 1.5 [0.7, 2.4] | 1.9 [1.1, 2.7] | 2.4 [1.6, 3.1] | 2.9 [2.2, 3.5] | 3.3 [2.6, 4.0] | 3.6 [2.8, 4.3] | 4.2 [3.0, 5.0] | 3.4 [2.4, 4.3] | |
| **75-79** | 0.9 [0.2, 2.1] | 1.5 [0.4, 2.3] | 2.0 [1.3, 2.7] | 2.5 [1.8, 3.2] | 3.0 [2.3, 3.7] | 3.4 [2.7, 4.1] | 3.6 [2.9, 4.3] | 4.3 [3.8, 5.0] | | |
| **80-84** | 0.9 [0.0, 1.8] | 1.4 [0.2, 2.4] | 2.1 [1.3, 2.8] | 2.6 [1.9, 3.3] | 3.0 [2.3, 3.6] | 3.4 [2.8, 4.2] | 4.0 [3.1, 4.6] | | | |
| **85-89** | 1.0 [0.0, 1.8] | 1.7 [0.6, 2.5] | 2.2 [1.3, 3.1] | 2.7 [1.9, 3.4] | 3.1 [2.4, 3.8] | 3.3 [2.6, 4.0] | 3.9 [3.4, 5.3] | | | |

### Adrenal gland (Total)

- **Male**

| | 1.3-1.4 | 1.4-1.5 | 1.5-1.6 | 1.6-1.7 | 1.7-1.8 | 1.8-1.9 | 1.9-2.0 | 2.0-2.1 | 2.1-2.2 | 2.2-2.3 |
|---|---|---|---|---|---|---|---|---|---|---|
| **20-24** | | 2.8 [1.7, 4.0] | 2.9 [1.7, 4.0] | 3.1 [2.5, 4.2] | 3.5 [2.9, 4.2] | 4.3 [2.9, 5.4] | 5.6 [4.5, 6.0] | | | |
| **25-29** | | | 3.2 [2.5, 4.2] | 3.6 [2.5, 4.7] | 4.3 [3.5, 5.8] | 5.4 [4.5, 7.7] | 5.6 [4.2, 6.2] | 6.6 [5.2, 8.2] | | |
| **30-34** | | | 3.8 [3.2, 4.5] | 4.3 [3.4, 5.3] | 4.8 [3.5, 5.9] | 4.9 [4.1, 5.9] | 6.2 [4.7, 7.4] | 6.4 [5.6, 7.7] | 7.4 [6.2, 9.0] | |
| **35-39** | | | 3.7 [2.5, 4.8] | 4.3 [3.5, 5.6] | 5.2 [4.0, 6.2] | 5.5 [4.1, 6.8] | 6.3 [5.3, 7.8] | 7.2 [6.2, 8.6] | 9.0 [7.2, 10.3] | |
| **40-44** | | 3.6 [2.4, 5.7] | 3.5 [2.7, 4.7] | 4.9 [3.6, 6.1] | 5.3 [4.2, 6.7] | 6.3 [5.3, 7.7] | 7.5 [6.1, 8.6] | 8.6 [6.5, 9.4] | 8.2 [7.2, 9.5] | |
| **45-49** | | 3.0 [2.4, 4.3] | 4.5 [3.5, 5.9] | 4.8 [3.5, 6.0] | 6.0 [4.8, 7.3] | 6.3 [5.3, 8.1] | 7.2 [5.8, 8.4] | 8.2 [7.0, 10.0] | 8.8 [7.3, 11.1] | 10.2 [9.5, 12.5] |
| **50-54** | | 4.1 [3.2, 5.3] | 4.7 [3.2, 6.1] | 5.6 [4.0, 6.9] | 6.3 [4.9, 7.5] | 6.8 [5.4, 8.2] | 7.8 [6.4, 9.0] | 8.7 [7.2, 10.0] | 9.3 [8.0, 10.7] | 10.4 [9.0, 12.5] |
| **55-59** | | 4.0 [3.0, 5.6] | 5.1 [3.6, 6.5] | 5.9 [4.6, 7.3] | 6.7 [5.4, 8.1] | 7.4 [6.0, 8.7] | 8.0 [6.5, 9.7] | 8.4 [7.4, 9.8] | 9.4 [8.5, 11.0] | 10.8 [8.4, 12.1] |
| **60-64** | 3.7 [3.1, 4.2] | 4.5 [3.0, 6.1] | 5.3 [3.8, 6.6] | 6.2 [4.8, 7.4] | 6.8 [5.6, 8.1] | 7.6 [6.5, 9.2] | 8.1 [6.9, 9.8] | 8.4 [7.2, 9.9] | 9.7 [7.2, 11.2] | 11.1 [8.5, 12.4] |
| **65-69** | 3.6 [2.0, 5.3] | 4.4 [3.3, 5.7] | 5.3 [3.7, 6.6] | 6.2 [5.1, 7.5] | 7.0 [5.7, 8.1] | 7.7 [6.3, 9.1] | 8.2 [6.8, 9.6] | 8.6 [7.2, 10.4] | 9.7 [8.5, 12.6] | 9.4 [7.9, 10.6] |
| **70-74** | 4.0 [2.0, 4.7] | 4.3 [3.1, 5.8] | 5.6 [4.1, 6.8] | 6.3 [5.0, 7.6] | 7.0 [5.8, 8.5] | 7.8 [6.6, 9.1] | 8.4 [6.9, 9.8] | 9.1 [7.8, 10.3] | 9.0 [7.9, 11.1] | |
| **75-79** | 4.0 [2.6, 5.1] | 4.7 [3.0, 6.3] | 5.7 [4.3, 6.9] | 6.6 [5.2, 7.9] | 7.0 [5.9, 8.4] | 7.6 [6.5, 9.1] | 8.2 [6.8, 9.7] | 9.1 [7.5, 10.6] | 9.2 [7.6, 11.7] | |
| **80-84** | 4.0 [2.4, 5.3] | 4.9 [3.2, 6.2] | 5.7 [4.5, 7.0] | 6.5 [5.3, 7.7] | 7.1 [5.8, 8.4] | 7.5 [6.4, 8.5] | 7.9 [6.7, 8.8] | 7.9 [7.0, 10.2] | | |
| **85-89** | 4.7 [2.0, 5.7] | 4.8 [3.8, 6.2] | 5.7 [4.3, 7.0] | 6.4 [5.2, 7.4] | 6.9 [5.7, 8.4] | 7.4 [6.2, 8.8] | 8.5 [6.6, 9.7] | | | |

- **Female**

| | 1.1-1.2 | 1.2-1.3 | 1.3-1.4 | 1.4-1.5 | 1.5-1.6 | 1.6-1.7 | 1.7-1.8 | 1.8-1.9 | 1.9-2.0 | 2.0-2.1 |
|---|---|---|---|---|---|---|---|---|---|---|
| **20-24** | | 1.6 [0.3, 2.8] | 2.2 [1.6, 2.6] | 2.1 [1.3, 2.6] | 2.4 [1.6, 3.0] | 2.7 [2.1, 3.6] | 3.4 [2.6, 4.2] | | | |
| **25-29** | | 1.7 [1.0, 3.2] | 2.3 [1.6, 2.9] | 2.2 [1.2, 2.9] | 3.0 [2.3, 3.8] | 3.1 [2.3, 3.6] | 4.8 [3.1, 5.5] | 4.1 [3.6, 5.1] | | |
| **30-34** | | 2.1 [1.5, 3.1] | 2.7 [1.9, 3.5] | 2.5 [1.7, 3.1] | 2.7 [2.0, 3.5] | 3.4 [2.7, 4.2] | 4.4 [3.1, 5.3] | 4.6 [3.9, 5.4] | 5.6 [4.7, 6.5] | |
| **35-39** | | 3.1 [1.9, 3.8] | 2.1 [1.3, 3.0] | 2.7 [2.1, 3.6] | 2.9 [2.2, 4.0] | 3.6 [3.0, 4.5] | 4.6 [3.6, 5.5] | 5.3 [4.5, 6.1] | 6.3 [4.9, 6.7] | |
| **40-44** | | 0.9 [0.5, 2.4] | 2.5 [1.6, 3.2] | 2.7 [1.8, 3.5] | 3.4 [2.5, 4.2] | 3.8 [2.8, 4.9] | 4.8 [3.8, 6.1] | 5.4 [4.3, 6.7] | 6.8 [5.6, 7.6] | 8.0 [7.3, 9.0] |
| **45-49** | | 2.0 [0.9, 2.7] | 2.5 [1.6, 3.2] | 3.0 [2.2, 3.9] | 3.7 [2.8, 4.6] | 4.1 [3.1, 5.2] | 5.1 [4.2, 6.3] | 6.3 [5.5, 7.4] | 7.2 [6.0, 8.6] | 6.7 [5.7, 7.9] |
| **50-54** | | 2.2 [1.2, 3.4] | 2.7 [1.8, 3.7] | 3.2 [2.3, 4.0] | 3.8 [2.9, 4.8] | 4.7 [3.6, 5.9] | 5.7 [4.4, 7.0] | 6.4 [5.3, 7.7] | 7.0 [5.9, 8.1] | 8.1 [6.5, 9.7] |
| **55-59** | | 1.8 [0.9, 2.9] | 2.6 [1.7, 3.6] | 3.3 [2.4, 4.4] | 4.1 [3.2, 5.2] | 4.9 [3.9, 6.1] | 6.1 [4.9, 7.5] | 6.6 [5.2, 7.8] | 7.6 [6.5, 8.7] | |
| **60-64** | 2.2 [0.7, 2.9] | 2.4 [1.2, 3.4] | 2.9 [1.9, 3.9] | 3.7 [2.8, 4.7] | 4.4 [3.4, 5.7] | 5.4 [4.3, 6.5] | 6.3 [5.1, 7.4] | 6.7 [5.5, 7.8] | 7.7 [7.0, 9.0] | 8.1 [7.0, 10.2] |
| **65-69** | 1.7 [0.7, 2.9] | 2.5 [1.4, 3.7] | 3.1 [2.0, 4.2] | 4.0 [2.9, 5.2] | 4.9 [3.8, 6.0] | 5.7 [4.6, 6.7] | 6.5 [5.6, 7.6] | 7.4 [6.1, 8.8] | 8.0 [7.0, 9.8] | |
| **70-74** | 1.9 [0.9, 3.4] | 2.8 [1.2, 3.9] | 3.4 [2.3, 4.7] | 4.3 [3.1, 5.5] | 5.2 [4.1, 6.3] | 6.1 [4.9, 7.3] | 6.4 [5.2, 7.7] | 7.2 [5.4, 8.7] | 6.3 [4.9, 8.5] | |
| **75-79** | 2.1 [1.0, 3.6] | 2.8 [1.5, 4.0] | 3.7 [2.5, 4.8] | 4.6 [3.4, 5.6] | 5.4 [4.3, 6.6] | 6.4 [4.9, 7.4] | 6.5 [5.7, 7.7] | 7.3 [6.0, 8.4] | | |
| **80-84** | 2.0 [1.1, 3.3] | 2.7 [1.4, 4.2] | 3.9 [2.6, 5.1] | 4.7 [3.7, 5.9] | 5.5 [4.3, 6.6] | 6.3 [5.5, 7.4] | 6.7 [5.9, 8.2] | | | |
| **85-89** | 2.0 [0.6, 3.5] | 3.0 [1.5, 4.4] | 4.1 [3.0, 5.3] | 5.0 [3.8, 5.9] | 5.7 [4.6, 6.8] | 5.9 [4.9, 7.1] | 6.9 [6.3, 9.1] | | | |

### Prostate

- **Male**

| | 1.3-1.4 | 1.4-1.5 | 1.5-1.6 | 1.6-1.7 | 1.7-1.8 | 1.8-1.9 | 1.9-2.0 | 2.0-2.1 | 2.1-2.2 | 2.2-2.3 |
|---|---|---|---|---|---|---|---|---|---|---|
| **20-24** | | 19.4 [15.3, 22.2] | 20.8 [16.2, 24.3] | 20.7 [17.2, 25.1] | 21.1 [15.7, 23.6] | 22.3 [19.5, 26.1] | 25.0 [20.7, 26.8] | | | |
| **25-29** | | | 20.6 [17.8, 23.2] | 19.9 [16.4, 24.0] | 22.4 [19.8, 24.2] | 23.5 [20.4, 25.0] | 23.7 [19.3, 25.8] | 20.7 [18.9, 26.1] | | |
| **30-34** | | | 19.5 [16.1, 23.3] | 22.0 [18.7, 24.1] | 24.0 [20.7, 27.1] | 23.1 [19.8, 25.8] | 25.6 [20.7, 27.9] | 26.9 [23.9, 28.3] | 28.8 [23.9, 33.3] | |
| **35-39** | | | 21.0 [16.6, 24.3] | 21.7 [19.9, 25.5] | 22.8 [19.9, 26.3] | 24.7 [21.3, 28.5] | 24.5 [20.9, 27.7] | 24.3 [18.7, 27.1] | 26.7 [24.5, 32.4] | |
| **40-44** | | 20.6 [12.9, 23.6] | 20.6 [16.8, 24.2] | 22.2 [19.7, 25.9] | 24.2 [21.0, 27.4] | 24.5 [20.6, 28.5] | 26.5 [22.8, 29.5] | 24.8 [22.1, 28.9] | 29.0 [25.0, 30.8] | |
| **45-49** | | 22.5 [14.0, 23.6] | 19.8 [17.6, 25.2] | 23.1 [19.0, 27.2] | 24.4 [20.5, 27.9] | 24.7 [21.3, 28.8] | 27.2 [22.9, 31.1] | 24.9 [22.1, 28.9] | 30.4 [25.2, 33.6] | 28.3 [25.2, 32.0] |
| **50-54** | | 19.8 [15.3, 21.5] | 21.0 [16.8, 24.6] | 23.1 [19.3, 26.8] | 24.8 [20.5, 28.4] | 25.3 [21.9, 28.9] | 26.9 [22.5, 30.2] | 27.6 [23.2, 31.2] | 27.4 [23.3, 33.5] | 31.5 [26.9, 34.7] |
| **55-59** | | 18.7 [14.6, 22.0] | 21.0 [17.7, 26.1] | 23.1 [18.9, 27.1] | 25.6 [21.6, 29.3] | 26.4 [22.4, 30.2] | 27.0 [22.5, 31.3] | 28.1 [23.8, 32.4] | 29.6 [25.2, 36.5] | 31.1 [27.8, 37.4] |
| **60-64** | 16.1 [13.7, 17.9] | 19.0 [14.9, 22.5] | 22.2 [17.1, 25.2] | 23.5 [19.0, 28.4] | 25.5 [21.2, 29.8] | 27.3 [22.3, 32.6] | 28.5 [23.0, 33.1] | 30.2 [24.6, 33.8] | 29.3 [24.6, 35.1] | 37.9 [24.2, 40.6] |
| **65-69** | 15.5 [12.2, 21.8] | 19.9 [15.3, 23.3] | 22.0 [17.0, 26.4] | 24.0 [18.9, 28.6] | 25.9 [21.2, 30.6] | 27.3 [21.0, 32.3] | 28.8 [23.0, 34.7] | 29.7 [23.7, 35.2] | 28.9 [24.4, 34.7] | 30.0 [28.5, 42.8] |
| **70-74** | 17.7 [9.9, 22.4] | 20.9 [15.9, 25.0] | 22.5 [17.4, 27.4] | 24.4 [18.7, 30.4] | 25.6 [20.2, 31.5] | 27.2 [22.1, 33.0] | 29.3 [23.3, 35.0] | 30.4 [24.1, 37.3] | 29.4 [24.8, 40.3] | |
| **75-79** | 17.5 [12.9, 23.1] | 21.1 [15.5, 26.1] | 22.4 [17.0, 27.1] | 24.4 [18.6, 30.7] | 26.4 [19.6, 32.2] | 28.5 [21.5, 34.9] | 28.4 [21.6, 37.1] | 29.4 [21.7, 40.1] | 33.8 [22.5, 41.4] | |
| **80-84** | 17.9 [10.7, 24.5] | 20.6 [15.1, 25.5] | 22.6 [16.9, 28.6] | 24.2 [18.0, 30.5] | 25.2 [19.3, 32.5] | 28.2 [21.3, 35.4] | 27.5 [19.5, 34.8] | 28.8 [23.7, 33.6] | | |
| **85-89** | 17.3 [10.9, 22.5] | 20.2 [13.6, 25.9] | 22.6 [15.1, 28.8] | 24.2 [17.1, 31.5] | 26.1 [18.5, 33.0] | 27.3 [19.2, 34.5] | 26.4 [23.1, 33.5] | | | |

### Iliopsoas muscle (Rt.)

- **Male**

| | 1.3-1.4 | 1.4-1.5 | 1.5-1.6 | 1.6-1.7 | 1.7-1.8 | 1.8-1.9 | 1.9-2.0 | 2.0-2.1 | 2.1-2.2 | 2.2-2.3 |
|---|---|---|---|---|---|---|---|---|---|---|
| **20-24** | | 223.0<br>[212.1, 262.9] | 278.7<br>[238.5, 316.0] | 301.6<br>[270.6, 341.3] | 313.8<br>[286.6, 350.2] | 348.8<br>[295.2, 389.0] | 361.9<br>[351.6, 396.3] | | | |
| **25-29** | | | 272.4<br>[220.5, 284.1] | 296.9<br>[273.7, 327.6] | 308.0<br>[267.1, 339.5] | 340.2<br>[309.3, 376.7] | 371.5<br>[347.0, 421.4] | 372.2<br>[302.3, 403.3] | | |
| **30-34** | | | 249.1<br>[225.4, 287.7] | 277.4<br>[242.6, 307.4] | 302.9<br>[269.9, 332.1] | 323.5<br>[299.6, 362.7] | 349.4<br>[315.5, 391.9] | 375.9<br>[319.1, 382.9] | 402.8<br>[370.5, 427.2] | |
| **35-39** | | | 256.2<br>[232.9, 289.4] | 279.2<br>[246.5, 307.6] | 297.6<br>[260.6, 322.6] | 329.4<br>[299.3, 357.0] | 354.3<br>[317.0, 374.9] | 344.5<br>[308.4, 376.2] | 380.1<br>[350.7, 461.4] | |
| **40-44** | | 192.3<br>[139.2, 212.2] | 238.0<br>[184.3, 278.4] | 268.8<br>[237.4, 296.6] | 292.0<br>[269.0, 326.2] | 321.7<br>[288.2, 349.3] | 339.2<br>[311.1, 365.7] | 369.0<br>[315.5, 401.6] | 371.1<br>[342.0, 399.2] | |
| **45-49** | | 204.8<br>[185.4, 236.0] | 220.1<br>[186.8, 256.5] | 261.1<br>[228.1, 285.8] | 287.1<br>[257.0, 317.1] | 306.8<br>[271.0, 338.1] | 331.2<br>[295.7, 360.8] | 340.6<br>[300.9, 371.6] | 368.5<br>[331.8, 402.3] | 368.2<br>[322.9, 406.9] |
| **50-54** | | 207.9<br>[180.3, 231.8] | 218.2<br>[187.5, 244.9] | 247.8<br>[219.2, 269.7] | 272.5<br>[244.8, 299.6] | 297.8<br>[265.7, 323.2] | 322.2<br>[283.8, 353.9] | 324.2<br>[291.1, 361.1] | 361.9<br>[314.8, 394.1] | 364.5<br>[317.9, 425.3] |
| **55-59** | | 191.6<br>[169.8, 228.8] | 220.6<br>[193.0, 238.6] | 241.2<br>[208.4, 265.7] | 260.8<br>[230.3, 287.7] | 288.0<br>[258.0, 316.2] | 301.5<br>[265.1, 330.7] | 325.1<br>[287.8, 353.2] | 346.2<br>[317.7, 373.7] | 346.2<br>[332.9, 383.1] |
| **60-64** | 169.5<br>[148.2, 177.5] | 191.4<br>[164.8, 208.8] | 205.8<br>[178.5, 231.2] | 237.6<br>[208.7, 260.7] | 258.4<br>[229.6, 282.1] | 275.2<br>[244.9, 302.6] | 294.8<br>[263.0, 320.1] | 304.0<br>[273.2, 337.5] | 334.7<br>[284.2, 365.9] | 334.4<br>[319.9, 366.9] |
| **65-69** | 156.9<br>[140.6, 190.1] | 185.0<br>[168.0, 212.2] | 201.6<br>[179.5, 229.2] | 231.5<br>[204.5, 258.6] | 246.9<br>[217.6, 274.2] | 260.4<br>[228.9, 288.8] | 282.4<br>[253.2, 315.2] | 300.0<br>[268.4, 328.1] | 311.3<br>[270.5, 346.3] | 350.7<br>[334.4, 394.1] |
| **70-74** | 159.4<br>[141.6, 181.8] | 176.8<br>[153.1, 197.1] | 201.1<br>[176.0, 222.8] | 218.8<br>[193.3, 243.2] | 238.1<br>[212.4, 262.8] | 252.7<br>[228.4, 282.2] | 272.1<br>[241.7, 303.1] | 303.4<br>[264.8, 332.4] | 311.3<br>[284.1, 341.2] | |
| **75-79** | 153.1<br>[134.5, 171.6] | 179.6<br>[158.3, 200.3] | 197.3<br>[170.1, 219.5] | 212.1<br>[185.5, 235.0] | 230.0<br>[205.1, 255.6] | 247.5<br>[219.4, 275.6] | 265.3<br>[238.4, 295.0] | 273.2<br>[250.9, 307.5] | 293.1<br>[262.4, 319.6] | |
| **80-84** | 156.5<br>[134.5, 172.3] | 174.1<br>[155.0, 192.5] | 190.6<br>[169.4, 212.9] | 209.4<br>[187.3, 231.5] | 225.5<br>[201.5, 247.9] | 243.1<br>[217.3, 270.2] | 263.4<br>[233.1, 282.9] | 268.2<br>[216.2, 307.5] | | |
| **85-89** | 167.0<br>[141.2, 179.7] | 172.0<br>[148.1, 189.4] | 190.1<br>[169.1, 209.0] | 203.0<br>[180.8, 228.1] | 213.4<br>[190.6, 237.6] | 231.8<br>[202.1, 258.2] | 253.8<br>[234.5, 277.6] | | | |

- **Female**

| | 1.1-1.2 | 1.2-1.3 | 1.3-1.4 | 1.4-1.5 | 1.5-1.6 | 1.6-1.7 | 1.7-1.8 | 1.8-1.9 | 1.9-2.0 | 2.0-2.1 |
|---|---|---|---|---|---|---|---|---|---|---|
| **20-24** | | 119.0<br>[114.4, 128.0] | 159.3<br>[138.1, 172.5] | 182.1<br>[167.3, 198.9] | 196.8<br>[177.4, 212.1] | 208.0<br>[186.3, 219.2] | 232.5<br>[197.7, 245.3] | | | |
| **25-29** | | 126.4<br>[112.0, 143.2] | 153.8<br>[137.8, 163.2] | 180.0<br>[161.0, 195.3] | 187.9<br>[169.9, 207.1] | 192.7<br>[171.6, 210.3] | 205.4<br>[190.3, 227.1] | 226.0<br>[206.7, 243.7] | | |
| **30-34** | | 110.8<br>[93.7, 131.0] | 159.0<br>[138.6, 166.3] | 173.1<br>[155.4, 189.1] | 186.7<br>[168.7, 201.1] | 205.6<br>[188.9, 219.5] | 200.0<br>[186.5, 221.0] | 214.4<br>[204.9, 234.4] | 275.6<br>[243.6, 308.8] | |
| **35-39** | | 126.4<br>[101.7, 138.2] | 154.6<br>[141.4, 171.6] | 176.7<br>[159.7, 192.1] | 185.3<br>[165.0, 205.6] | 197.4<br>[182.1, 216.9] | 222.2<br>[195.6, 241.1] | 225.3<br>[201.2, 235.8] | 213.6<br>[183.0, 238.1] | |
| **40-44** | | 127.8<br>[104.5, 142.6] | 153.2<br>[138.3, 174.4] | 171.6<br>[152.9, 188.5] | 182.4<br>[167.6, 198.2] | 197.0<br>[178.1, 214.9] | 209.8<br>[190.5, 224.2] | 226.1<br>[201.0, 247.3] | 229.6<br>[207.5, 254.8] | 242.3<br>[224.3, 266.4] |
| **45-49** | | 138.6<br>[123.3, 147.6] | 151.0<br>[135.4, 166.9] | 163.2<br>[147.4, 181.5] | 177.0<br>[160.2, 194.4] | 189.6<br>[170.8, 202.7] | 201.2<br>[185.2, 218.9] | 211.3<br>[193.1, 237.6] | 216.5<br>[202.9, 244.8] | 240.6<br>[218.9, 247.8] |
| **50-54** | | 125.9<br>[106.7, 139.7] | 144.6<br>[124.3, 161.9] | 160.2<br>[144.2, 177.5] | 173.2<br>[155.3, 188.2] | 183.5<br>[164.8, 200.3] | 191.6<br>[174.6, 213.9] | 199.6<br>[177.7, 230.0] | 221.5<br>[204.8, 241.9] | 223.6<br>[203.7, 235.4] |
| **55-59** | | 126.4<br>[105.2, 143.7] | 145.7<br>[127.5, 159.7] | 156.7<br>[138.8, 172.5] | 167.7<br>[149.5, 181.6] | 176.7<br>[160.9, 193.7] | 190.5<br>[168.9, 210.6] | 205.1<br>[178.8, 230.3] | 213.9<br>[194.6, 241.0] | |
| **60-64** | 117.5<br>[113.6, 140.0] | 129.2<br>[113.7, 148.0] | 140.7<br>[123.4, 152.6] | 153.8<br>[138.2, 169.0] | 163.2<br>[144.1, 181.0] | 173.7<br>[155.2, 194.0] | 180.2<br>[168.3, 199.7] | 197.7<br>[181.5, 216.4] | 207.6<br>[190.2, 224.2] | 224.0<br>[192.7, 234.9] |
| **65-69** | 114.6<br>[98.4, 127.9] | 122.9<br>[109.1, 136.9] | 137.9<br>[124.8, 153.8] | 149.0<br>[131.0, 164.2] | 160.2<br>[141.9, 176.3] | 169.8<br>[151.1, 188.4] | 182.2<br>[164.8, 201.1] | 190.3<br>[176.3, 213.0] | 197.3<br>[185.8, 214.9] | |
| **70-74** | 109.8<br>[101.1, 125.1] | 122.6<br>[107.3, 136.2] | 134.9<br>[117.6, 150.6] | 146.7<br>[128.3, 161.4] | 156.0<br>[138.9, 173.4] | 165.5<br>[147.7, 184.8] | 182.4<br>[160.1, 198.8] | 190.5<br>[162.3, 207.7] | 183.5<br>[166.3, 218.5] | |
| **75-79** | 116.2<br>[101.0, 127.8] | 125.1<br>[109.3, 139.5] | 132.1<br>[115.2, 147.0] | 142.1<br>[125.9, 158.7] | 154.5<br>[134.4, 170.5] | 164.9<br>[149.0, 182.3] | 179.5<br>[161.4, 192.1] | 172.2<br>[150.7, 188.2] | | |
| **80-84** | 114.6<br>[92.5, 133.6] | 124.7<br>[111.0, 141.2] | 130.3<br>[115.2, 146.5] | 143.0<br>[126.3, 158.6] | 155.1<br>[136.1, 169.5] | 159.9<br>[142.3, 180.0] | 164.1<br>[144.5, 195.5] | | | |
| **85-89** | 105.5<br>[86.5, 123.8] | 123.3<br>[107.6, 138.3] | 130.9<br>[111.4, 146.2] | 141.2<br>[123.9, 157.6] | 149.7<br>[129.9, 165.2] | 162.0<br>[141.8, 175.1] | 167.8<br>[162.1, 181.9] | | | |

### Iliopsoas muscle (Lt.)

#### - Male

| | 1.3-1.4 | 1.4-1.5 | 1.5-1.6 | 1.6-1.7 | 1.7-1.8 | 1.8-1.9 | 1.9-2.0 | 2.0-2.1 | 2.1-2.2 | 2.2-2.3 |
|---|---|---|---|---|---|---|---|---|---|---|
| **20-24** | | 230.3 [210.7, 278.3] | 282.1 [241.2, 305.2] | 318.5 [271.5, 352.1] | 326.5 [289.7, 358.9] | 355.6 [299.7, 403.6] | 383.4 [352.0, 413.5] | | | |
| **25-29** | | | 270.0 [228.5, 282.0] | 304.5 [273.0, 323.2] | 317.7 [273.2, 346.4] | 344.3 [313.3, 380.7] | 398.6 [370.6, 418.6] | 393.9 [329.8, 436.1] | | |
| **30-34** | | | 248.1 [221.3, 279.0] | 282.5 [248.2, 308.3] | 306.4 [277.4, 341.1] | 329.9 [300.7, 375.7] | 358.4 [331.9, 400.6] | 375.9 [336.8, 397.0] | 412.0 [385.1, 437.7] | |
| **35-39** | | | 261.4 [229.2, 300.0] | 287.9 [273.8, 318.8] | 297.0 [263.3, 331.4] | 334.3 [313.4, 366.7] | 365.5 [324.2, 385.1] | 342.9 [315.4, 392.9] | 409.2 [359.3, 466.9] | |
| **40-44** | | 202.4 [139.8, 230.7] | 240.4 [184.8, 279.7] | 273.3 [239.4, 298.5] | 295.5 [265.8, 328.8] | 331.7 [297.4, 357.0] | 350.5 [315.9, 381.8] | 363.8 [334.8, 415.2] | 375.0 [353.9, 408.4] | |
| **45-49** | | 203.0 [184.6, 245.2] | 228.9 [191.8, 251.7] | 262.8 [230.2, 290.2] | 287.7 [260.4, 320.2] | 314.7 [280.9, 345.7] | 329.7 [300.5, 368.8] | 346.3 [308.0, 384.5] | 392.6 [339.7, 425.6] | 392.3 [351.9, 434.8] |
| **50-54** | | 206.4 [179.7, 227.3] | 222.1 [189.0, 246.3] | 253.4 [222.5, 279.5] | 276.6 [248.2, 304.6] | 306.5 [277.2, 333.8] | 333.4 [295.1, 367.4] | 337.4 [295.3, 376.1] | 374.0 [332.9, 408.9] | 388.0 [341.9, 414.9] |
| **55-59** | | 190.3 [170.5, 212.2] | 217.6 [186.9, 243.8] | 244.5 [213.4, 269.2] | 265.9 [239.7, 295.5] | 296.8 [267.0, 326.6] | 313.9 [274.3, 346.9] | 333.3 [297.7, 360.4] | 368.1 [325.2, 391.2] | 374.1 [353.7, 428.1] |
| **60-64** | 172.7 [122.9, 198.6] | 194.1 [175.1, 209.8] | 205.6 [177.8, 233.0] | 240.8 [214.3, 269.6] | 265.9 [237.8, 293.3] | 284.5 [253.6, 317.9] | 308.7 [274.4, 336.6] | 320.9 [290.6, 352.6] | 346.5 [309.3, 379.5] | 362.5 [325.6, 386.5] |
| **65-69** | 157.1 [141.6, 185.2] | 184.9 [168.0, 206.0] | 207.1 [179.2, 232.8] | 234.9 [209.1, 261.4] | 256.9 [227.1, 285.6] | 273.2 [239.1, 302.1] | 300.6 [263.6, 328.4] | 318.8 [291.3, 346.4] | 330.2 [292.2, 373.0] | 367.1 [337.3, 372.3] |
| **70-74** | 155.5 [134.6, 182.6] | 178.7 [153.8, 201.8] | 202.9 [180.3, 224.6] | 226.7 [198.1, 251.7] | 247.6 [221.8, 273.9] | 267.2 [238.6, 296.6] | 286.0 [255.1, 316.0] | 316.5 [268.6, 344.2] | 339.9 [286.5, 359.6] | |
| **75-79** | 153.0 [132.4, 170.1] | 181.2 [154.6, 202.5] | 200.6 [174.9, 221.3] | 219.7 [191.8, 244.1] | 240.1 [213.1, 264.4] | 261.6 [230.7, 290.6] | 281.3 [252.2, 310.1] | 285.9 [263.3, 316.6] | 298.4 [284.7, 339.2] | |
| **80-84** | 151.6 [132.6, 175.8] | 175.2 [155.8, 196.6] | 197.5 [175.0, 221.3] | 218.6 [193.2, 242.6] | 238.0 [210.0, 261.5] | 257.4 [229.2, 281.4] | 274.3 [248.2, 303.1] | 295.2 [255.8, 337.5] | | |
| **85-89** | 158.6 [139.9, 181.0] | 170.5 [149.0, 192.2] | 192.8 [171.0, 211.8] | 210.2 [188.8, 235.1] | 225.8 [201.5, 253.9] | 246.3 [215.8, 277.3] | 269.7 [247.2, 286.4] | | | |

#### - Female

| | 1.1-1.2 | 1.2-1.3 | 1.3-1.4 | 1.4-1.5 | 1.5-1.6 | 1.6-1.7 | 1.7-1.8 | 1.8-1.9 | 1.9-2.0 | 2.0-2.1 |
|---|---|---|---|---|---|---|---|---|---|---|
| **20-24** | | 121.3 [106.3, 132.5] | 165.3 [145.3, 175.2] | 186.0 [166.0, 200.5] | 207.7 [186.1, 221.6] | 208.5 [196.6, 218.0] | 233.7 [209.7, 251.3] | | | |
| **25-29** | | 128.4 [117.7, 147.0] | 158.7 [146.7, 167.6] | 184.9 [164.4, 197.3] | 185.2 [174.3, 213.9] | 199.4 [180.6, 218.1] | 221.6 [196.3, 233.2] | 231.0 [212.4, 252.3] | | |
| **30-34** | | 120.7 [102.4, 136.4] | 159.0 [145.0, 170.6] | 173.8 [159.1, 193.5] | 188.5 [168.5, 204.3] | 204.7 [195.0, 222.2] | 217.3 [201.2, 244.5] | 227.4 [204.7, 236.7] | 279.8 [268.3, 305.7] | |
| **35-39** | | 129.8 [107.5, 137.6] | 161.9 [141.9, 173.3] | 177.6 [163.9, 193.4] | 190.8 [171.9, 207.7] | 203.7 [191.7, 221.3] | 222.5 [204.0, 249.6] | 236.9 [217.0, 251.2] | 225.6 [182.3, 251.7] | |
| **40-44** | | 124.7 [109.0, 146.5] | 157.7 [136.8, 180.0] | 174.9 [153.9, 191.9] | 189.1 [171.6, 203.2] | 202.3 [182.7, 222.2] | 213.6 [195.4, 233.7] | 227.5 [212.4, 253.1] | 237.2 [212.6, 268.7] | 256.5 [233.3, 290.7] |
| **45-49** | | 133.4 [115.0, 147.3] | 154.2 [134.1, 169.7] | 166.4 [148.7, 185.5] | 181.8 [163.9, 197.3] | 194.9 [176.7, 213.0] | 208.7 [190.8, 228.7] | 219.9 [199.5, 240.9] | 231.0 [211.0, 266.3] | 255.0 [224.5, 277.1] |
| **50-54** | | 132.1 [112.1, 139.1] | 144.0 [125.6, 168.2] | 163.3 [145.5, 178.4] | 177.6 [162.4, 194.7] | 189.8 [171.6, 207.0] | 201.1 [181.3, 218.8] | 210.6 [186.2, 235.7] | 239.9 [221.3, 262.2] | 243.0 [208.3, 254.8] |
| **55-59** | | 125.1 [108.9, 143.0] | 146.1 [130.8, 162.1] | 160.9 [143.2, 177.5] | 172.6 [154.8, 189.5] | 181.5 [163.9, 200.8] | 200.0 [177.0, 221.1] | 215.0 [193.4, 238.6] | 229.3 [216.5, 238.5] | |
| **60-64** | 113.9 [98.1, 125.1] | 130.1 [110.6, 149.8] | 143.3 [126.3, 158.4] | 156.3 [141.1, 173.9] | 168.8 [148.9, 186.6] | 181.4 [164.3, 202.8] | 192.2 [174.7, 208.8] | 209.0 [184.6, 229.2] | 214.6 [194.5, 231.3] | 228.8 [213.1, 255.5] |
| **65-69** | 116.4 [100.9, 124.0] | 122.3 [102.9, 137.9] | 142.9 [126.0, 156.1] | 152.2 [133.1, 169.7] | 165.3 [145.9, 181.9] | 177.8 [156.9, 195.6] | 192.6 [175.8, 215.0] | 207.2 [178.1, 220.0] | 216.6 [203.2, 230.1] | |
| **70-74** | 117.9 [99.2, 123.8] | 125.2 [107.4, 139.5] | 137.6 [118.9, 153.3] | 149.6 [132.2, 166.9] | 161.1 [144.2, 180.2] | 176.9 [155.2, 194.7] | 190.5 [164.9, 210.5] | 200.0 [170.0, 216.9] | 196.6 [182.8, 225.3] | |
| **75-79** | 113.2 [88.2, 129.5] | 128.8 [110.2, 143.8] | 136.2 [119.4, 150.7] | 149.2 [131.8, 165.4] | 160.1 [141.3, 178.0] | 174.2 [154.2, 193.6] | 189.9 [166.0, 206.1] | 187.6 [163.5, 205.5] | | |
| **80-84** | 112.3 [94.9, 123.6] | 126.1 [111.9, 141.8] | 135.6 [120.1, 151.7] | 150.6 [132.2, 164.3] | 160.4 [139.4, 178.3] | 172.5 [154.0, 192.5] | 178.4 [153.2, 201.0] | | | |
| **85-89** | 107.1 [85.6, 126.2] | 124.7 [108.1, 137.1] | 134.8 [116.7, 150.9] | 146.3 [128.0, 163.8] | 158.5 [136.8, 177.5] | 176.3 [151.8, 189.1] | 182.4 [172.4, 209.6] | | | |

### Iliopsoas muscle (Total)

#### - Male

| | 1.3-1.4 | 1.4-1.5 | 1.5-1.6 | 1.6-1.7 | 1.7-1.8 | 1.8-1.9 | 1.9-2.0 | 2.0-2.1 | 2.1-2.2 | 2.2-2.3 |
|---|---|---|---|---|---|---|---|---|---|---|
| **20-24** | | 456.6<br>[434.1, 549.9] | 546.0<br>[488.8, 627.7] | 618.9<br>[547.3, 689.2] | 639.0<br>[584.8, 709.0] | 706.0<br>[595.2, 802.0] | 741.7<br>[704.4, 809.8] | | | |
| **25-29** | | | 546.4<br>[448.8, 561.1] | 598.4<br>[547.4, 653.1] | 626.2<br>[550.5, 688.3] | 684.7<br>[623.3, 763.6] | 767.3<br>[719.6, 847.1] | 763.1<br>[634.5, 835.8] | | |
| **30-34** | | | 484.4<br>[455.1, 573.8] | 557.5<br>[488.6, 605.3] | 604.3<br>[549.9, 672.7] | 654.2<br>[594.5, 733.6] | 704.4<br>[641.1, 794.7] | 750.0<br>[658.1, 778.8] | 832.4<br>[755.5, 864.9] | |
| **35-39** | | | 517.6<br>[463.7, 593.2] | 563.8<br>[523.6, 624.1] | 589.7<br>[524.5, 655.8] | 662.7<br>[607.7, 719.1] | 719.6<br>[651.0, 762.7] | 679.3<br>[629.3, 758.5] | 789.3<br>[686.7, 929.0] | |
| **40-44** | | 394.8<br>[278.9, 442.9] | 480.2<br>[370.5, 562.3] | 542.1<br>[476.2, 601.6] | 589.6<br>[539.1, 649.9] | 652.3<br>[584.4, 708.0] | 693.0<br>[625.8, 744.8] | 730.3<br>[650.9, 816.0] | 748.2<br>[693.7, 801.9] | |
| **45-49** | | 411.8<br>[376.9, 481.2] | 437.0<br>[377.4, 511.9] | 531.4<br>[451.8, 571.5] | 576.5<br>[514.7, 638.0] | 619.6<br>[555.0, 682.0] | 662.1<br>[597.3, 726.8] | 687.4<br>[610.2, 749.3] | 757.2<br>[674.4, 829.3] | 780.4<br>[679.8, 833.0] |
| **50-54** | | 424.1<br>[361.7, 460.8] | 447.3<br>[391.0, 487.5] | 501.8<br>[448.1, 548.1] | 547.6<br>[493.0, 601.1] | 602.0<br>[546.8, 655.5] | 654.8<br>[576.7, 719.7] | 657.2<br>[586.4, 738.6] | 740.1<br>[651.3, 804.1] | 755.7<br>[659.8, 836.6] |
| **55-59** | | 377.4<br>[344.1, 422.4] | 438.7<br>[377.3, 482.3] | 489.8<br>[424.5, 532.0] | 525.1<br>[472.1, 578.6] | 582.3<br>[525.2, 642.6] | 614.6<br>[542.0, 675.3] | 656.1<br>[585.9, 715.2] | 717.5<br>[636.9, 761.8] | 719.4<br>[678.5, 809.7] |
| **60-64** | 342.1<br>[287.0, 399.3] | 385.8<br>[330.4, 416.6] | 413.0<br>[357.3, 461.0] | 477.2<br>[425.6, 527.8] | 523.0<br>[469.2, 576.9] | 559.7<br>[500.9, 621.8] | 605.0<br>[539.2, 657.5] | 626.6<br>[566.5, 691.3] | 675.0<br>[598.7, 749.3] | 695.4<br>[658.8, 755.9] |
| **65-69** | 321.5<br>[291.4, 379.8] | 366.7<br>[335.8, 416.0] | 410.0<br>[364.5, 459.4] | 467.5<br>[412.5, 516.1] | 503.9<br>[451.1, 558.6] | 533.2<br>[471.3, 587.9] | 584.6<br>[519.7, 643.8] | 620.8<br>[556.8, 666.7] | 638.8<br>[558.6, 726.7] | 710.9<br>[662.7, 777.9] |
| **70-74** | 316.6<br>[285.7, 357.1] | 350.1<br>[307.3, 395.2] | 403.3<br>[358.7, 444.0] | 444.3<br>[395.0, 492.8] | 484.3<br>[435.0, 535.4] | 520.8<br>[466.5, 575.6] | 557.7<br>[496.5, 613.9] | 617.6<br>[535.8, 673.2] | 651.6<br>[596.0, 692.8] | |
| **75-79** | 308.7<br>[273.7, 344.0] | 357.1<br>[317.2, 400.7] | 398.7<br>[347.8, 439.5] | 430.8<br>[378.7, 478.6] | 470.2<br>[418.6, 519.1] | 508.2<br>[452.3, 566.7] | 545.5<br>[488.7, 601.3] | 555.4<br>[519.6, 614.6] | 603.1<br>[548.4, 661.1] | |
| **80-84** | 302.9<br>[274.5, 352.8] | 348.9<br>[312.5, 389.6] | 389.5<br>[346.8, 433.0] | 428.9<br>[381.9, 471.2] | 460.8<br>[412.3, 507.3] | 500.4<br>[448.8, 548.7] | 533.1<br>[486.8, 588.0] | 562.8<br>[460.3, 635.4] | | |
| **85-89** | 316.3<br>[288.3, 360.0] | 339.2<br>[297.6, 381.3] | 384.4<br>[341.0, 421.6] | 414.1<br>[368.3, 460.1] | 438.2<br>[395.2, 487.5] | 477.7<br>[424.0, 532.7] | 527.9<br>[484.6, 567.3] | | | |

#### - Female

| | 1.1-1.2 | 1.2-1.3 | 1.3-1.4 | 1.4-1.5 | 1.5-1.6 | 1.6-1.7 | 1.7-1.8 | 1.8-1.9 | 1.9-2.0 | 2.0-2.1 |
|---|---|---|---|---|---|---|---|---|---|---|
| **20-24** | | 240.2<br>[220.7, 261.2] | 324.4<br>[289.3, 345.1] | 370.2<br>[332.8, 400.8] | 403.5<br>[364.6, 428.6] | 417.9<br>[380.5, 429.1] | 466.1<br>[407.8, 498.3] | | | |
| **25-29** | | 254.6<br>[225.3, 292.3] | 312.6<br>[286.3, 327.7] | 366.4<br>[324.2, 392.4] | 372.0<br>[341.7, 423.5] | 393.8<br>[346.5, 426.8] | 427.1<br>[377.0, 463.5] | 455.5<br>[425.5, 499.5] | | |
| **30-34** | | 229.9<br>[197.0, 266.1] | 317.5<br>[289.5, 338.6] | 347.8<br>[312.0, 375.7] | 372.6<br>[336.4, 407.6] | 414.4<br>[389.7, 436.6] | 416.4<br>[390.5, 469.0] | 446.1<br>[404.2, 471.1] | 565.2<br>[519.2, 609.5] | |
| **35-39** | | 255.7<br>[214.5, 284.6] | 316.3<br>[287.0, 344.1] | 353.9<br>[321.3, 380.6] | 374.8<br>[338.9, 409.7] | 399.6<br>[374.7, 438.1] | 441.2<br>[396.4, 486.7] | 461.8<br>[417.5, 484.7] | 431.5<br>[361.4, 489.7] | |
| **40-44** | | 253.8<br>[218.6, 289.1] | 309.5<br>[278.8, 355.4] | 345.3<br>[304.3, 379.0] | 372.6<br>[340.2, 400.2] | 399.2<br>[362.1, 432.3] | 427.0<br>[383.9, 452.1] | 452.0<br>[414.1, 502.9] | 467.4<br>[437.0, 517.2] | 487.6<br>[452.6, 556.4] |
| **45-49** | | 270.0<br>[237.6, 296.6] | 304.2<br>[272.2, 333.5] | 328.3<br>[294.9, 366.7] | 361.5<br>[325.6, 389.8] | 386.4<br>[346.3, 414.7] | 409.3<br>[379.5, 445.4] | 431.6<br>[391.5, 476.1] | 447.5<br>[415.7, 511.6] | 494.9<br>[444.0, 525.3] |
| **50-54** | | 263.9<br>[217.2, 277.5] | 288.5<br>[251.5, 326.3] | 322.6<br>[292.4, 354.5] | 351.4<br>[318.5, 383.2] | 374.3<br>[336.7, 407.6] | 391.9<br>[355.9, 433.1] | 415.8<br>[364.6, 465.6] | 462.4<br>[428.3, 507.5] | 469.5<br>[416.2, 481.5] |
| **55-59** | | 253.2<br>[217.0, 284.1] | 291.7<br>[262.1, 320.0] | 317.5<br>[284.4, 348.2] | 339.8<br>[305.9, 371.5] | 357.8<br>[322.9, 393.2] | 391.1<br>[349.1, 427.9] | 423.6<br>[381.1, 466.6] | 440.0<br>[406.7, 473.7] | |
| **60-64** | 229.7<br>[212.6, 260.1] | 253.3<br>[225.9, 295.0] | 285.1<br>[251.3, 310.7] | 310.4<br>[282.5, 342.1] | 331.7<br>[294.6, 367.6] | 354.1<br>[321.8, 398.7] | 371.2<br>[347.4, 406.0] | 406.3<br>[369.7, 443.8] | 423.2<br>[384.0, 455.7] | 457.9<br>[389.7, 491.2] |
| **65-69** | 228.4<br>[194.2, 258.8] | 240.3<br>[220.3, 274.2] | 280.9<br>[251.5, 309.5] | 301.9<br>[263.0, 333.6] | 324.3<br>[290.8, 355.7] | 348.3<br>[308.6, 384.2] | 378.1<br>[340.3, 414.2] | 396.9<br>[357.9, 423.3] | 417.2<br>[390.2, 442.6] | |
| **70-74** | 222.3<br>[200.8, 250.7] | 248.5<br>[214.1, 272.2] | 273.4<br>[239.1, 302.5] | 296.4<br>[262.1, 327.7] | 317.5<br>[286.7, 352.0] | 342.1<br>[303.1, 378.6] | 374.4<br>[324.3, 402.0] | 384.5<br>[337.5, 420.3] | 376.7<br>[353.8, 443.8] | |
| **75-79** | 228.0<br>[187.4, 263.8] | 253.5<br>[220.2, 283.7] | 267.2<br>[235.0, 295.4] | 290.3<br>[259.8, 321.9] | 315.5<br>[276.8, 346.2] | 340.9<br>[304.7, 376.7] | 365.7<br>[331.0, 402.6] | 357.6<br>[326.1, 382.1] | | |
| **80-84** | 224.1<br>[190.0, 259.9] | 250.0<br>[225.0, 281.2] | 265.7<br>[236.4, 295.5] | 292.3<br>[261.5, 321.3] | 313.9<br>[276.5, 346.1] | 333.0<br>[299.1, 374.9] | 339.6<br>[304.0, 391.9] | | | |
| **85-89** | 209.8<br>[178.1, 254.3] | 247.2<br>[220.3, 270.2] | 266.4<br>[229.6, 294.6] | 286.6<br>[251.3, 317.7] | 304.7<br>[272.1, 341.1] | 340.6<br>[294.3, 364.3] | 347.9<br>[341.5, 392.7] | | | |
